# Social inequalities in COVID-19 deaths by area-level income: patterns over time and the mediating role of vaccination in a population of 11.2 million people in Ontario, Canada

**DOI:** 10.1101/2024.01.15.24301331

**Authors:** Linwei Wang, Sarah Swayze, Korryn Bodner, Andrew Calzavara, Sean P. Harrigan, Arjumand Siddiqi, Stefan D. Baral, Peter C. Austin, Janet Smylie, Maria Koh, Hind Sbihi, Beate Sander, Jeffrey C. Kwong, Sharmistha Mishra

## Abstract

**Importance:** Social inequalities in COVID-19 deaths were evident early in the pandemic. Less is known about how vaccination may have influenced inequalities in COVID-19 deaths.

**Objectives:** To examine patterns in COVID-19 deaths by area-level income over time and to examine the impact of vaccination on inequality patterns in COVID-19 deaths.

**Design, setting, and participants:** Population-based retrospective cohort study including community-living individuals aged ≥18 years residing in Ontario, Canada, as of March 1, 2020 who were followed through to January 30, 2022 (five pandemic waves).

**Exposure:** Area-level income derived from the 2016 Census at the level of dissemination area categorized into quintiles. Vaccination defined as receiving ≥ 1 dose of Johnson-Johnson vaccine or ≥ 2 doses of other vaccines.

**Main outcome measures:** COVID-19 death defined as death within 30 days following, or 7 days prior to a positive SARS-CoV-2 PCR test. Cause-specific hazard models were used to examine the relationship between income and COVID-19 deaths in each wave. We used regression-based causal mediation analyses to examine the impact of vaccination in the relationship between income and COVID-19 deaths during waves four and five.

**Results:** Of 11,248,572 adults, 7044 (0.063%) experienced a COVID-19 death. After accounting for demographics, baseline health, and area-level social determinants of health, inequalities in COVID-19 deaths by income persisted over time (adjusted hazard ratios (aHR) [95% confidence intervals] comparing lowest-income vs. highest-income quintiles were 1.37[0.98-1.92] for wave one, 1.21[0.99-1.48] for wave two, 1.55[1.22-1.96] for wave three, and 1.57[1.15-2.15] for waves four and five). Of 11,122,816 adults alive by the start of wave four, 7,534,259(67.7%) were vaccinated, with lower odds of vaccination in the lowest-income compared to highest-income quintiles (0.71[0.70-0.71]). This inequality in vaccination accounted for 57.9%[21.9%-94.0%] of inequalities in COVID-19 deaths between individuals in the lowest-income vs. highest-income quintiles.

**Conclusions:** Inequalities by income persisted in COVID-19 deaths over time. Efforts are needed to address both vaccination gaps and residual heightened risks associated with lower income to improve health equity in COVID-19 outcomes.

**Summary box:** *Section 1: What is already known on this topic:*

- Emerging data suggest social inequalities in COVID-19 deaths might have persisted over time, but existing studies were limited by their ecological design and/or inability to account for potential confounders.
- Vaccination has contributed to reducing COVID-19 deaths but there were social inequalities in vaccination coverage.
- The impact of inequalities in vaccination on inequalities in COVID-19 deaths has not yet been well-studied.

*Section 2: What this study adds:*

- Across five pandemic waves (2020-2021) in Ontario, Canada, COVID-19 deaths remained higher in individuals living in lower-income neighbourhoods, even after accounting for individual-level demographics and baseline health, and other area-level social determinants of health.
- During later waves (following the vaccination roll-out), over half (57.9%) of the inequalities in COVID-19 deaths between individuals living in the lowest and highest income neighbourhoods could be attributed to differential vaccination coverage by income. This means that if vaccine equality was achieved, inequalities in deaths would persist but be reduced.
- Addressing vaccination gaps, as well as addressing the residual heightened risks of COVID-19 associated with lower income could improve health equity in COVID-19 outcomes.

## Introduction

Globally, the number of confirmed COVID-19 cases has surpassed 770 million with over 6.95 million deaths(1). The burden of COVID-19 has been disproportionately borne by individuals and communities experiencing socioeconomic disadvantage, leading to regional inequalities in COVID-19 outcomes within and between countries(2–7). Social inequalities in COVID-19 deaths were evident during the early pandemic period across the world(2–8). Data are emerging to suggest social inequalities in COVID-19 deaths might have persisted into later pandemic waves in the UK(9), the US(10) and Canada(11). However, these studies were limited by their ecological design(10,11) and/or inability to account for potential confounders including comorbidities(9,11).

Global COVID-19 vaccination programmes are estimated to have prevented 19.8 million deaths worldwide during the first year of roll-out(12), but vaccination coverage has been consistently unequal by socioeconomic factors(13). Social inequalities in vaccination coverage were evident between countries(e.g., countries with lower income had lower vaccination coverage(14)) and within countries (e.g., individuals living in neighbourhoods with greater socioeconomic disadvantage had lower likelihood of vaccination(13,15)), driven by systemic forces that shaped unequal vaccine access(16–18).

There is a limited, but growing body of empirical evidence on the role of unequal vaccination coverage on inequalities in COVID-19 deaths(10,19). An ecological study in the US found that social inequalities in vaccination coverage between regions contributed to social inequalities in COVID-19 case fatality between regions(10), highlighting the potential mediating role of vaccination in social inequalities in COVID-19 deaths. Another ecological study in the UK showed that lower area-level income was associated with greater area-level COVID-19 mortality(19). The study also found that the magnitude of inequality was smaller in neighbourhoods with higher vaccination coverage, highlighting the potential for effect modification by vaccination in social inequalities in COVID-19 deaths(19). Thus, there is a need for individual-level studies to examine the impact of vaccination as it was rolled-out, on social inequalities in COVID-19 deaths.

Leveraging population-based individual-level data among 11.2 million adults in Ontario, Canada, we sought to examine changes over time in inequalities in COVID-19 deaths by area-level income, and to examine the role of vaccination on inequalities in COVID-19 deaths. First, we quantified the patterns in COVID-19 deaths by area-level income across five pandemic waves (March 1, 2020 to January 30, 2022) accounting for individual-level demographics and baseline health, and area-level social determinants of health (SDOH). Second, we quantified patterns in vaccination by area-level income by the start of wave four (August 1, 2020), and examined the role of vaccination, as a potential effect modifier and/or mediator, in the relationship between area-level income and COVID-19 deaths during waves four and five.

## METHODS

### Study design and subjects

We conducted a population-based retrospective cohort study of community-dwelling adults in Ontario, Canada. Individuals aged ≥18 years residing in Ontario as of March 1, 2020, and with a provincial health insurance were identified using Ontario’s Registered Persons Database (RPDB) and followed through to January 30, 2022. We excluded individuals aged >105 years, long-term care home residents, individuals with non-Ontario postal codes, and those without healthcare system contact for >3 years (if aged 65+) or >9 years (if aged<65); we additionally excluded individuals with missing covariates (**Appendix Figure 1**).

### Measures

Our primary outcome was COVID-19 death, defined as death within 30 days following or 7 days prior to a positive SARS-CoV-2 test, consistent with a prior study(3). Test result and date were determined using records in the Ontario COVID-19 surveillance database (Public Health Case and Contact Management Solution (CCM)) and the Ontario Laboratories Information System. Date of death was determined using CCM and RPDB, which capture over 99.3 % of COVID-19 deaths in Ontario(3). We ascertained COVID-19 deaths between March 1, 2020 and January 30, 2022, with a positive test date between March 1, 2020 and December 31, 2021 to allow for a minimum 30 days follow-up.

We classified the study period into five pandemic waves based on the positive test date, and in line with regional pandemic characteristics(20). Epidemic curves across the five waves, the dates correspond to each wave, and cumulative vaccination coverage over time are shown in **Figure 1**. We combined wave four and wave five (referred to as ‘waves four&five) in model analyses to increase analytic power given small numbers of COVID-19 deaths in each wave.

**Figure 1.**
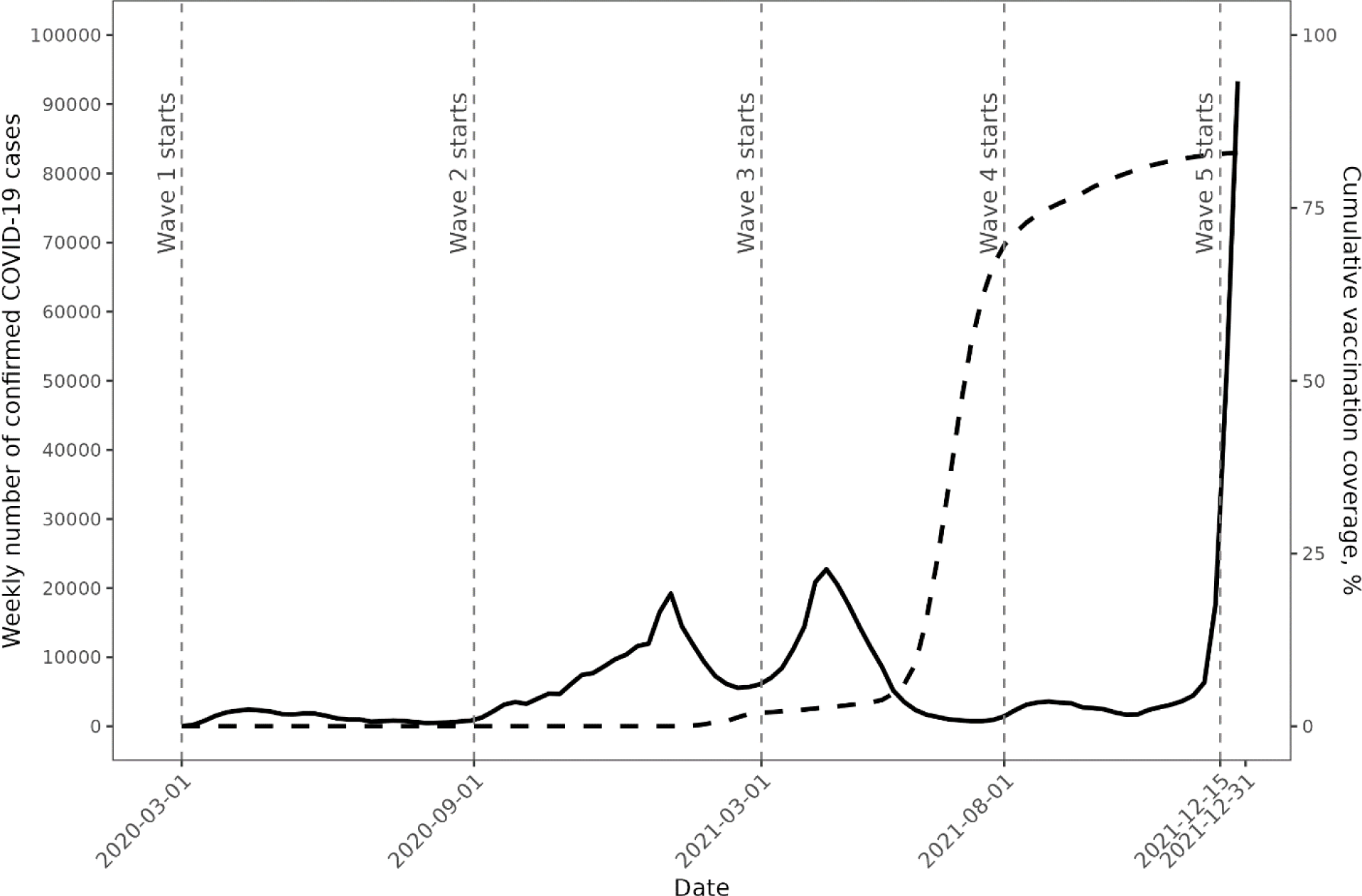
Epidemic curves (solid line reflects weekly numbers of confirmed COVID-19 cases) and cumulative vaccination coverage (dash line) across five pandemic waves between March 1, 2020 and December 31, 2021 in Ontario, Canada. Dates refer to the start of an epidemiological week. Vaccination defined as receiving ≥ 1 dose of Johnson-Johnson vaccine or ≥ 2 doses of other vaccines.

Our primary exposure was area-level income derived from the 2016 census at the level of dissemination area (DA), the smallest geographic unit (representing 400-700 residents) for which census data are reported(21). We ranked DAs at the city-level and categorized them into quintiles. For example, a DA being in income quintile 1 means it is among the lowest 20% of DAs in its city by median household income.

Our primary mediator variable of interest was vaccination status (yes vs. no) defined as receiving ≥ 1 dose of Johnson-Johnson vaccine or ≥ 2 doses of other vaccines, derived from Ontario’s COVID-19 vaccination registry(details in **Appendix Text 1**).

We considered other covariates given their potential as exposure-outcome confounders and/or mediator-outcome confounders(3,4,13,22) (**Figure 2A**). Given data availability, these covariates included individual-level demographics (age; sex; immigration status based on the Immigration, Refugees and Citizenship Canada Permanent Resident Database(23,24); living in rural vs. urban area; public health unit of residence), baseline health (composite comorbidity measure using the ACG^®^ System Aggregated Diagnosis Groups generated using the Johns Hopkins ACG® System Version 10(25); past 3-year hospital admission; past year outpatient physician visits), prior infection (yes vs. no), and other area-level SDOH (proportion with diploma or higher educational attainment; proportion essential workers; proportion racially-minoritized groups; proportion apartment buildings; proportion high density housing; average household size). Detailed definitions and data sources for these variables are shown in **Table 1 footnotes.**

**Figure 2.**
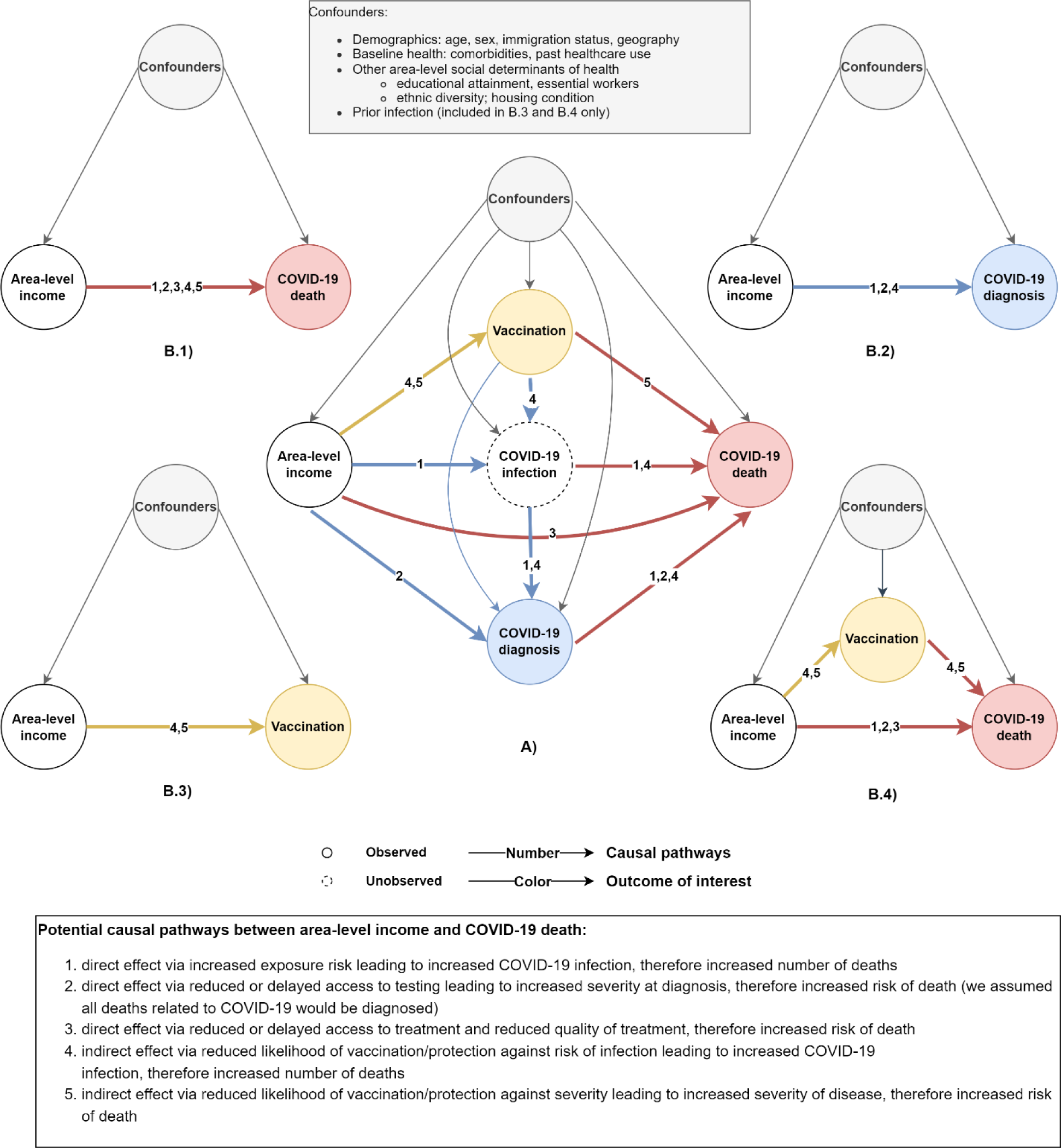
Directed acyclic graphs depicting A) summary of potential causal pathways between area-level income and COVID-19 deaths (we define pathways via vaccination as indirect, otherwise, direct), and B) proposed analyses adjusting for confounders estimating B.1) total effect of area-level income on COVID-19 deaths; B.2) total effect of area-level income on COVID-19 diagnosws; B.3) total effect of area-level income on vaccination status; and B.4) direct and indirect (mediated through vaccination) effect of area-level income on COVID-19 deaths.

**Table 1.** Characteristics of community-dwelling adults in Ontario as of March 1, 2020 stratified by area-level income quintiles (N=1,1248,572).

| Characteristics |  | Area-level median household income quintiles (Q1=Lowest-income quintile)<sup>a,b</sup> |  |  |  |  |
| --- | --- | --- | --- | --- | --- | --- |
|  |  | Q1<br>N=2,194,815 | Q2<br>N=2,231,228 | Q3<br>N=2,280,638 | Q4<br>N=2,269,199 | Q5<br>N=2,272,692 |
| COVID-19 death<sup>c</sup> |  | 2122 (0.097%) | 1684 (0.075%) | 1363 (0.060%) | 1004 (0.044%) | 871 (0.038%) |
| Age category |  |  |  |  |  |  |
|  | 20-34 | 674,633 (30.7%) | 644,437 (28.9%) | 639,087 (28.0%) | 612,066 (27.0%) | 577,562 (25.4%) |
|  | 35-49 | 526,561 (24.0%) | 532,274 (23.9%) | 574,860 (25.2%) | 594,588 (26.2%) | 562,492 (24.8%) |
|  | 50-64 | 537,377 (24.5%) | 562,619 (25.2%) | 586,966 (25.7%) | 602,777 (26.6%) | 641,230 (28.2%) |
|  | 65-74 | 256,471 (11.7%) | 279,891 (12.5%) | 282,796 (12.4%) | 276,709 (12.2%) | 298,268 (13.1%) |
|  | 75-84 | 140,858 (6.4%) | 151,066 (6.8%) | 144,211 (6.3%) | 136,078 (6.0%) | 143,974 (6.3%) |
|  | 85+ | 58,915 (2.7%) | 60,941 (2.7%) | 52,718 (2.3%) | 46,981 (2.1%) | 49,166 (2.2%) |
| Male |  | 1,060,313 (48.3%) | 1,083,403 (48.6%) | 1,116,067 (48.9%) | 1,111,752 (49.0%) | 1,114,429 (49.0%) |
| Residing in a rural area<sup>d</sup> |  | 222,499 (10.1%) | 226,820 (10.2%) | 223,477 (9.8%) | 230,715 (10.2%) | 237,399 (10.4%) |
| Immigration status<sup>e</sup> |  |  |  |  |  |  |
|  | Long-term resident | 1,576,783 (71.8%) | 1,701,199 (76.2%) | 1,761,391 (77.2%) | 1,806,881 (79.6%) | 1,932,511 (85.0%) |
|  | Long-term immigrant | 484,753 (22.1%) | 433,674 (19.4%) | 438,518 (19.2%) | 397,370 (17.5%) | 292,741 (12.9%) |
|  | Recent (within 5 years) immigrant | 133,279 (6.1%) | 96,355 (4.3%) | 80,729 (3.5%) | 64,948 (2.9%) | 47,440 (2.1%) |
| Aggregated Diagnosis Groups<sup>f</sup> |  |  |  |  |  |  |
|  | 0 | 256,107 (11.7%) | 240,963 (10.8%) | 230,998 (10.1%) | 219,681 (9.7%) | 220,719 (9.7%) |
|  | 1-2 | 425,185 (19.4%) | 441,881 (19.8%) | 454,369 (19.9%) | 461,696 (20.3%) | 470,558 (20.7%) |
|  | 3-4 | 460,305 (21.0%) | 487,282 (21.8%) | 512,184 (22.5%) | 520,638 (22.9%) | 527,436 (23.2%) |
|  | 5-6 | 396,352 (18.1%) | 415,754 (18.6%) | 434,863 (19.1%) | 437,828 (19.3%) | 437,614 (19.3%) |
|  | 7+ | 656,866 (29.9%) | 645,348 (28.9%) | 648,224 (28.4%) | 629,356 (27.7%) | 616,365 (27.1%) |
| Hospital admission, past 3 years |  |  |  |  |  |  |
|  | 0 times | 1,870,952 (85.2%) | 1,927,879 (86.4%) | 1,980,778 (86.9%) | 1,984,343 (87.4%) | 2,001,914 (88.1%) |
|  | Once | 217,763 (9.9%) | 211,946 (9.5%) | 214,214 (9.4%) | 206,809 (9.1%) | 196,962 (8.7%) |
|  | Twice | 61,026 (2.8%) | 55,101 (2.5%) | 53,333 (2.3%) | 49,769 (2.2%) | 4,7350 (2.1%) |
|  | Three times or more | 45,074 (2.1%) | 36,302 (1.6%) | 32,313 (1.4%) | 28,278 (1.2%) | 264,66 (1.2%) |
| Outpatient physician visits, past year |  |  |  |  |  |  |
|  | 0-1 times | 730,273 (33.3%) | 716,329 (32.1%) | 717,362 (31.5%) | 709,167 (31.3%) | 713,001 (31.4%) |
|  | 2-4 times | 563,370 (25.7%) | 600,462 (26.9%) | 634,090 (27.8%) | 651,578 (28.7%) | 668,168 (29.4%) |
|  | 5-8 times | 434,005 (19.8%) | 455,002 (20.4%) | 473,212 (20.7%) | 474,597 (20.9%) | 473,776 (20.8%) |
|  | 9-14 times | 281,217 (12.8%) | 285,120 (12.8%) | 288,022 (12.6%) | 278,077 (12.3%) | 269,777 (11.9%) |
|  | 15 times or more | 185,950 (8.5%) | 174,315 (7.8%) | 167,952 (7.4%) | 155,780 (6.9%) | 147,970 (6.5%) |
| Educational attainment quintile (1=Lowest)<sup>a,g</sup> |  |  |  |  |  |  |
|  | 1 | 983,358 (44.8%) | 537,946 (24.1%) | 249,371 (10.9%) | 92,773 (4.1%) | 29,420 (1.3%) |
|  | 2 | 589,663 (26.9%) | 646,515 (29.0%) | 526,964 (23.1%) | 295,138 (13.0%) | 126,588 (5.6%) |
|  | 3 | 327,869 (14.9%) | 450,727 (20.2%) | 673,994 (29.6%) | 596,976 (26.3%) | 323,015 (14.2%) |
|  | 4 | 169,587 (7.7%) | 371,973 (16.7%) | 513,292 (22.5%) | 712,452 (31.4%) | 670,934 (29.5%) |
|  | 5 | 124,338 (5.7%) | 224,067 (10.0%) | 317,017 (13.9%) | 571,860 (25.2%) | 1,122,735 (49.4%) |
| Proportion essential workers quintile (1=Lowest)<sup>a,h</sup> |  |  |  |  |  |  |
|  | 1 | 127,561 (5.8%) | 253,151 (11.3%) | 377,604 (16.6%) | 535,585 (23.6%) | 1,093,115 (48.1%) |
|  | 2 | 216,121 (9.8%) | 386,266 (17.3%) | 477,337 (20.9%) | 770,519 (34.0%) | 662,379 (29.1%) |
|  | 3 | 356,241 (16.2%) | 461,993 (20.7%) | 577,151 (25.3%) | 511,656 (22.5%) | 338,508 (14.9%) |
|  | 4 | 552,795 (25.2%) | 552,504 (24.8%) | 563,572 (24.7%) | 340,174 (15.0%) | 143,864 (6.3%) |
|  | 5 | 942,097 (42.9%) | 577,314 (25.9%) | 284,974 (12.5%) | 111,265 (4.9%) | 34,826 (1.5%) |
| Proportion racially-minoritised quintile(1=Lowest)<sup>a,i</sup> |  |  |  |  |  |  |
|  | 1 | 279,103 (12.7%) | 327,935 (14.7%) | 416,887 (18.3%) | 400,328 (17.6%) | 325,650 (14.3%) |
|  | 2 | 277,463 (12.6%) | 370,618 (16.6%) | 359,571 (15.8%) | 380,955 (16.8%) | 488,678 (21.5%) |
|  | 3 | 297,255 (13.5%) | 365,057 (16.4%) | 346,677 (15.2%) | 402,145 (17.7%) | 605,123 (26.6%) |
|  | 4 | 392,499 (17.9%) | 428,067 (19.2%) | 482,436 (21.2%) | 555,335 (24.5%) | 602,481 (26.5%) |
|  | 5 | 948,495 (43.2%) | 739,551 (33.1%) | 675,067 (29.6%) | 530,436 (23.4%) | 250,760 (11.0%) |
| Proportion apartment buildings (1=Lowest)<sup>a,j</sup> | 1 | 252,283 (11.5%) | 906,097 (40.6%) | 1,517,122 (66.5%) | 1,842,461 (81.2%) | 1,908,598 (84.0%) |
|  | 2 | 447,483 (20.4%) | 626,594 (28.1%) | 488,640 (21.4%) | 264,403 (11.7%) | 221,414 (9.7%) |
|  | 3 | 1,495,049 (68.1%) | 698,537 (31.3%) | 274,876 (12.1%) | 162,335 (7.2%) | 142,680 (6.3%) |
| Average household size quintile (1=Lowest)<sup>a,k</sup> | 1 | 964,195 (43.9%) | 589,097 (26.4%) | 280,769 (12.3%) | 161,762 (7.1%) | 135,383 (6.0%) |
|  | 2 | 459,829 (21.0%) | 586,066 (26.3%) | 463,777 (20.3%) | 295,561 (13.0%) | 172,838 (7.6%) |
|  | 3 | 198,744 (9.1%) | 273,057 (12.2%) | 362,191 (15.9%) | 378,511 (16.7%) | 320,635 (14.1%) |
|  | 4 | 313,709 (14.3%) | 345,761 (15.5%) | 517,598 (22.7%) | 654,512 (28.8%) | 829,702 (36.5%) |
|  | 5 | 258,338 (11.8%) | 437,247 (19.6%) | 656,303 (28.8%) | 778,853 (34.3%) | 814,134 (35.8%) |
| Proportion high-density housing (1=Lowest)<sup>a,l</sup> | 1 | 300,492 (13.7%) | 513,834 (23.0%) | 741,894 (32.5%) | 964,003 (42.5%) | 1,387,054 (61.0%) |
|  | 2 | 353,488 (16.1%) | 464,887 (20.8%) | 531,676 (23.3%) | 652,045 (28.7%) | 496,239 (21.8%) |
|  | 3 | 344,062 (15.7%) | 469,729 (21.1%) | 609,939 (26.7%) | 493,387 (21.7%) | 311,631 (13.7%) |
|  | 4 | 1,196,773 (54.5%) | 782,778 (35.1%) | 397,129 (17.4%) | 159,764 (7.0%) | 77,768 (3.4%) |
| Vaccination status by the start of wave 4 (August 1, 2021)<sup>m</sup> |  |  |  |  |  |  |
| Fully vaccinated with booster |  | 63 (0.003%) | 61 (0.003%) | 89 (0.004%) | 83 (0.004%) | 145 (0.006%) |
| Fully vaccinated without booster |  | 1,296,266 (59.1%) | 1,444,411 (64.7%) | 1,532,756 (67.2%) | 1,584,421 (69.8%) | 1,685,027 (74.1%) |
| Partially vaccinated |  | 265,256 (12.1%) | 245,661 (11.0%) | 241,202 (10.6%) | 222,792 (9.8%) | 185,256 (8.2%) |
| Unvaccinated |  | 633,230 (28.9%) | 541,095 (24.3%) | 506,591 (22.2%) | 461,903 (20.4%) | 402,264 (17.7%) |
| Vaccination status by the start of wave 5 (December 15, 2021)<sup>m</sup> |  |  |  |  |  |  |
| Fully vaccinated with booster |  | 137,878 (6.3%) | 157,866 (7.1%) | 167,956 (7.4%) | 182,253 (8.0%) | 247,493 (10.9%) |
| Fully vaccinated without booster |  | 1,573,459 (71.7%) | 1,660,237 (74.4%) | 1,728,131 (75.8%) | 1,736,535 (76.5%) | 1,708,954 (75.2%) |
| Partially vaccinated |  | 58,048 (2.6%) | 45,845 (2.1%) | 41,609 (1.8%) | 366,65 (1.6%) | 31,330 (1.4%) |
| Unvaccinated |  | 425,430 (19.4%) | 367,280 (16.5%) | 342,942 (15.0%) | 313,746 (13.8%) | 284,915 (12.5%) |
| Prior infection<sup>n</sup> |  | 1121 (0.052%) | 901 (0.041%) | 854 (0.038%) | 751 (0.033%) | 682 (0.030%) |

<sup>a</sup>Measured at the level of the census dissemination area.
<sup>b</sup>Dissemination areas were ranked within each city by their median household income to create quintiles; we ranked within city instead of within the entire province to take the cost of living into account; a dissemination area being in quintile 1 means it is among the lowest 20% of dissemination areas in its city by median household income.
<sup>c</sup>Death within 30 days following or 7 days prior to a lab-confirmed positive SARS-CoV-2 test.
<sup>d</sup>Rural defined as being located outside the commuting zone of a city with a population greater than 10000.
<sup>e</sup>Individual-level data from the Immigration, Refugees and Citizenship Canada (IRCC)'s Permanent Resident Database.
<sup>f</sup>Calculated using Johns Hopkins ACG® System Version 10 with 2-year look-back; databases used : Discharge Abstract Database, National Ambulatory Care Reporting System, Ontario Health Insurance Plan billings, Ontario Drug Benefits Plan, Continuing Care Reporting System, Canadian Organ Replacement Registry, Ontario Cancer Registry.
<sup>g</sup>1<sup>st</sup> quintile represents areas with 17.1-94.3% of people aged 25-64 years without a diploma; 2<sup>nd</sup>, 11.4-17.1%; 3<sup>rd</sup>, 7.5%-11.4%; 4<sup>th</sup>, 4.1-7.5%; and 5<sup>th</sup>, 0-4.1%.
<sup>h</sup>1<sup>st</sup> quintile represents 0%-32.5% of working people in the area who self-identified as working in an essential job, including sales, trades, manufacturing, and agriculture; 2<sup>nd</sup> quintile, 32.5%-42.3% of people; 3<sup>rd</sup> quintile, 42.3%-49.8% of people; 4<sup>th</sup> quintile, 50.0%-57.5% of people; and 5<sup>th</sup> quintile, 57.5%-114.3% of people.
<sup>i</sup>1<sup>st</sup> quintile represents 0%-2.2% of people in the area who self-identified as part of a racially-minoritised group(s); 2<sup>nd</sup> quintile, 2.2%-7.5% of people; 3<sup>rd</sup> quintile, 7.5%-18.7% of people; 4<sup>th</sup> quintile, 18.7%-43.5% of people; and 5<sup>th</sup> quintile, 43.5%-100% of people; racially-minoritised groups defined as people who self-identify as non-White and non-Indigenous;
<sup>j</sup>1<sup>st</sup> category, 0%-7.3% of buildings in the area are apartment buildings; 2<sup>nd</sup> category, 7.4%-37.7% are apartment buildings; and 3<sup>rd</sup> category, 37.7%-100% are apartment buildings; the high frequency of zeros permitted the creation of only 3 categories (i.e., the lower 3 quintiles combined, and the fourth and fifth quintiles).
<sup>k</sup>1<sup>st</sup> quintile represents 0-2.1 people/dwelling; 2<sup>nd</sup>, 2.2-2.4 people/dwelling; 3<sup>rd</sup>, 2.5-2.6 people/dwelling; 4<sup>th</sup>, 2.7-3 people/dwelling; and 5<sup>th</sup>, 3.1-5.7 people/dwelling.
<sup>l</sup>1<sup>st</sup> category represents 0-2.6% of households are considered high-density housing; 2<sup>nd</sup> y, 2.7-5.2%; 3<sup>rd</sup>, 5.3-8.7%; 4<sup>th</sup>, >8.7%; the high frequency of zeros permitted the creation of only 4 categories (the lower 2 quintiles combined); 'housing density' refers to whether a private household is living in suitable accommodations according to the National Occupancy Standard; that is, whether the dwelling has enough bedrooms for the size and composition of the household.
<sup>m</sup>Details on definition of vaccination status in **Appendix text 1**.
<sup>n</sup>Defined as whether an individual had a positive SARS-CoV-2 test between March 1, 2020 and August 1, 2021 (the start of wave 4).

All data sets were linked using unique encoded identifiers and analyzed at ICES(26). ICES is an independent, non-profit research institute whose legal status under Ontario’s health information privacy law allows it to collect and analyze health care and demographic data, without consent, for health system evaluation and improvement. Data use is authorized under section 45 of Ontario’s Personal Health Information Protection Act and does not require review by a Research Ethics Board. Patients or the public were not involved in the design, or conduct, or reporting, or dissemination plans of our research

### Statistical analyses

#### Descriptive analyses

We examined the number of individuals at risk, and the number of COVID-19 diagnoses and deaths for each wave. We compared the characteristics at baseline, and vaccination status at the start of each wave, by income quintiles.

#### Regression analyses for objective 1: social inequality patterns over time (across five waves)

To examine changes over time in COVID-19 deaths by income, we first fitted separate cause-specific hazard models(27,28) using SAS 9.4(29) to examine the relationship between area-level income and COVID-19 deaths during each wave, treating deaths without a positive SARS-CoV-2 test as competing risk events (**Figure 2B.1**). We fitted unadjusted models and models adjusted for demographics, baseline health, and area-level SDOH. We did not adjust for vaccination status nor prior infection to capture the total effect of area-level income on COVID-19 deaths including pathways mediated by vaccination and prior infection, and to ensure comparability across waves. We subsequently fitted a model among the full sample with interaction terms between income and wave (wave treated as a continuous variable) to test the trend in the magnitude of the associations between income and COVID-19 deaths over time. We accounted for clustering by DA using a robust sandwich covariance matrix(30).

#### Sensitivity analyses for objective 1

We repeated the above analyses for our secondary outcome (COVID-19 diagnoses) to examine the changes over time in COVID-19 diagnosis patterns by income (**Figure 2B.2**).

To assess the impact of COVID-19 variants on social inequality patterns(31), we repeated the above analyses for variant-specific outcomes using waves four&five as an example. We focused on Delta-related and Omicron-related deaths and diagnoses as they were dominant during waves four&five. The classification of COVID-19 variants was based on a combination of whole genome sequencing data when available, mutation screening results, and the date of diagnosis (details in **Appendix Text 2**).

#### Regression analyses for objective 2: the impact of vaccination in social inequality patterns (waves four&five)

We examined the impact of vaccination by the start of wave four on social inequality patterns during waves four&five. We restricted objective 2 analyses to waves four&five because the vaccination coverage was zero or very low for earlier waves (**Table 1**).

We hypothesized that vaccination status could be a mediator for the relationship between income and COVID-19 deaths such that area-level income affects individual’s likelihood of vaccination, leading to differential protection against risk of infection and severity therefore differential risk of COVID-19 deaths (**Figure 2A**). We also hypothesized that vaccination could modify the relationship between income and COVID-19 deaths such that the magnitude of social inequalities in COVID-19 deaths could differ between subgroups with different vaccination status (**Appendix text 3**).

We first fitted logistic regression models to examine the relationship between income and the odds of vaccination by the start of wave four (**Figure 2B.3**). We then employed regression-based causal mediation analyses(32,33) in R 4.1.2(34) using the ‘CMAverse’ package(35) to first examine the interaction between income and vaccination (effect modification), and then to estimate the total, direct, and indirect effects of income on COVID-19 deaths and the proportion of total effects attributable to mediation by vaccination(**Figure 2B.4**). We adjusted for individual-level demographics and baseline health, area-level SDOH, and prior infection.

#### Sensitivity analyses for objective 2

We performed quantitative bias analyses for unmeasured confounding using an E-value approach(36,37) for the mediation analyses. The E-value is described as the minimum strength of association required between an unmeasured confounder with both exposure and outcome to nullify the observed association(36). E-values from 1.0-1.5, 1.5-3.0, and >3.0 suggest weak, moderate and strong unmeasured confounding, respectively(36).

To contextualize the magnitude of the vaccination impact as a mediator given multiple pathways between income and COVID-19 deaths, we conducted two separate mediation analyses to examine the total, direct and indirect effect of income on COVID-19 deaths, treating our measures of area-level essential worker and housing density as a mediator, respectively in each model(**Appendix Figure 2**).

## Results

Of 11,248,572 adults (median age 48 years [interquartile range: 33-62 years]) included, 613,338 (5.5%) tested positive for SARS-CoV-2, of whom 7,044 (1.1%) experienced a COVID-19 death. COVID-19 deaths were disproportionately concentrated among individuals in the lowest-income vs. highest-income quintiles (0.097% vs. 0.038%) (**Table 1**). Compared to the highest-income quintile, individuals in the lowest-income quintile were more likely to be recent immigrants (6.1% vs. 2.1%), to be hospitalized in the past three years (14.8% vs. 11.9%), and to be living in neighbourhoods characterized by lower educational attainment, higher proportion essential workers, higher proportion racially minoritized groups, and higher density housing (**Table 1**).

A total of 11,248,572; 11,207,019; 11,160,968; and 11,122,816 individuals were at risk for SARS-CoV-2 infection by the start wave one, wave two, wave three, and waves four&five; of whom, 30,356; 191,945; 176,438; 214,599 tested positive for SARS-CoV-2, resulting in 955, 2860, 2185, and 1044 COVID-19 deaths, during wave one, wave two, wave three, and waves four&five, respectively. Of the 1,044 COVID-19 deaths during waves four&five, 651(62.4%) were Delta-related, 321(30.7%) were Omicron-related, and the reminder due to other or unknown variants.

By the start of wave four, 7,534,259 (67.7%) individuals were vaccinated. Individuals in the lowest-income quintile were less likely to be vaccinated than individuals in the highest-income quintile (59.1% vs 74.1%) (**Table 1**).

### Patterns across pandemic waves

After adjusting for demographics, baseline health, and area-level SDOH, living in lower-income neighbourhoods was consistently associated with an increased hazard of COVID-19 death across waves (**Figure 3A.1**). Compared to the highest-income quintile, the lowest-income quintile was associated with 1.37 times [95% confidence interval (CI): 0.98-1.92] hazard of COVID-19 death during wave one, 1.21 times [0.99-1.48] during wave two, 1.55 times [1.22-1.96] during wave three, and 1.57 times [1.15-2.15] during waves four&five (**Figure 3A.1; Appendix Table 1**). A trend test revealed an increasing trend over time in the magnitude of the association (p-value=0.02).

**Figure 3.**
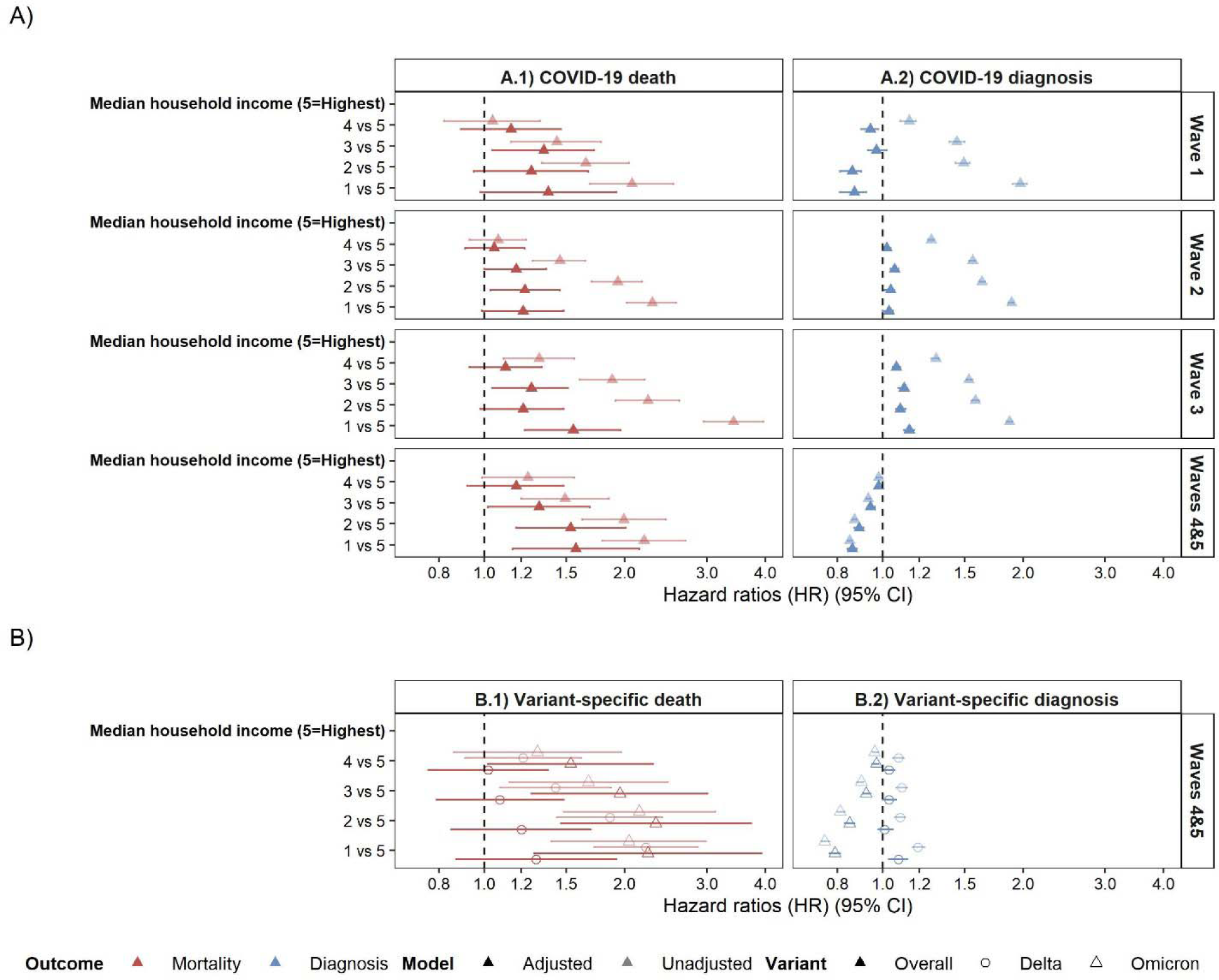
Patterns by area-level income quintiles in A) overall COVID-19 deaths and diagnoses stratified by pandemic wave; and B) variant-specific deaths and diagnoses during waves 4&5. Waves were classified based on the date of diagnosis, and categorized into wave 1: Mar 1-Aug 31, 2020; wave 2: Sep 1, 2020–Feb 28, 2021; wave 3: Mar 1–Jul 31, 2021; and waves 4&5: Aug 1-Dec 31, 2021. Adjusted analyses adjusted for individual-level demographics, baseline health, and other area-level social determinants of health.

Patterns in COVID-19 diagnoses by income were less consistent across waves (**Figure 3A.2**). Compared to the highest-income quintile, the lowest-income quintile was associated with an increased hazard of diagnosis during waves two and three, and a decreased hazard of diagnosis during waves one and four&five. The adjusted hazard ratios (aHR) for waves one to four&five were as follows: 0.87 [0.81-0.92], 1.03 [1.00-1.05], 1.14 [1.11-1.17], and 0.86 [0.84-0.88] (**Figure 3A.2; Appendix Table 1**).

### Sensitivity analyses: variant-specific outcomes during waves four&five

Compared to the highest-income quintile, the lowest-income quintile was associated with an increased hazard of Delta-diagnosis (aHR: 1.08 [1.03-1.13]), and an increased hazard of Delta-related death (1.29 [0.87-1.92]); in contrast, the lowest-income quintile was associated with a decreased hazard of Omicron-diagnosis (0.79 [0.77-0.81]), and an increased hazard of Omicron-related death (2.24 [1.28-3.93]) (**Figure 3B; Appendix Table 2**).

### Mediating and effect modifying role of vaccination during waves four&five

Lower income was independently associated with a decreased odds of vaccination by the start of wave four (**Figure 4A; Appendix Table 3**). For example, the adjusted odds ratio (aOR) comparing the lowest-income vs. highest-income quintiles was 0.71 [0.70-0.71].

**Figure 4.**
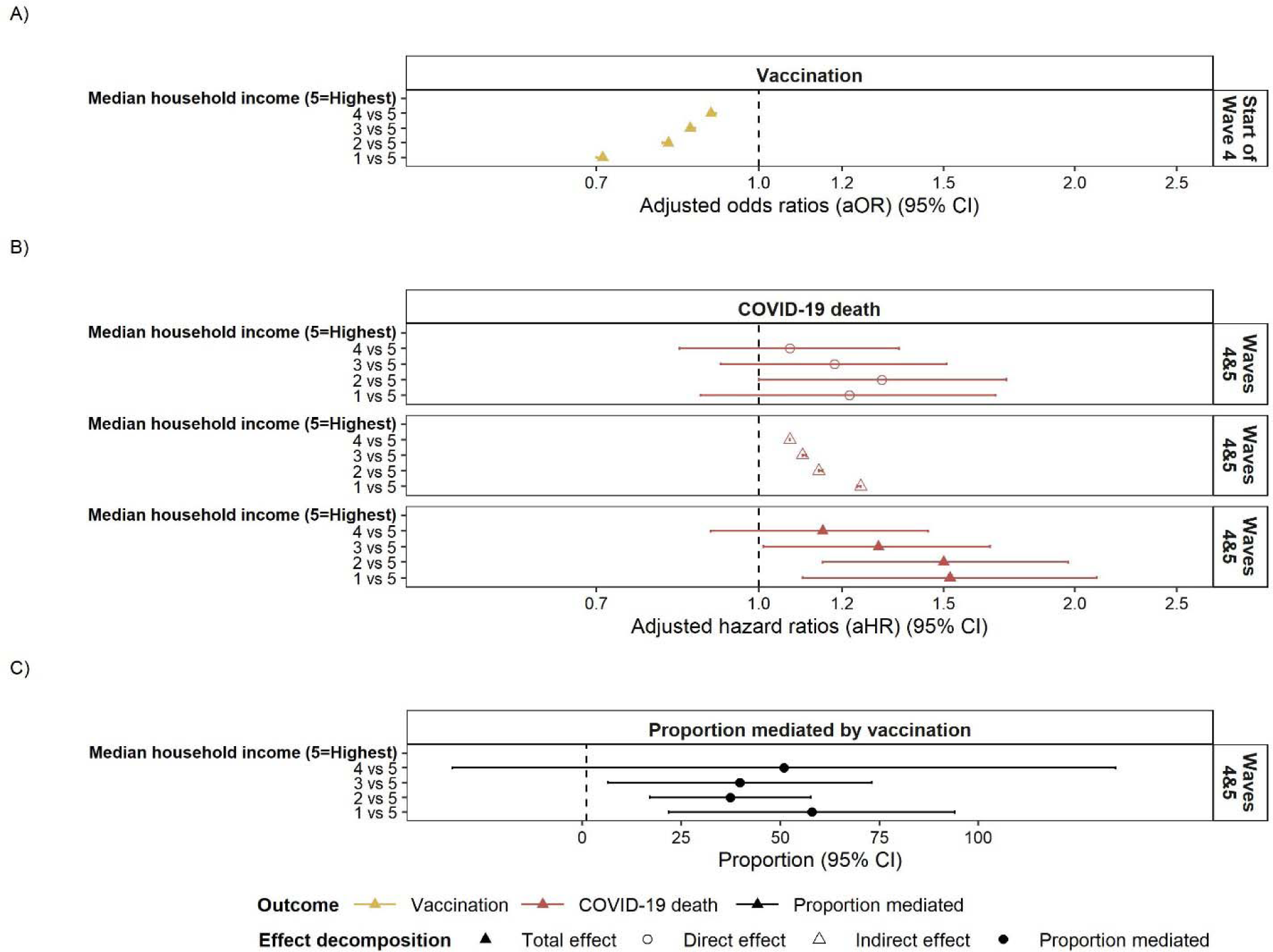
Results of causal mediation analyses including A) the relationship between area-level income (exposure) and vaccination (mediator); B) the relationship between area-level income (exposure) and COVID-19 deaths (outcome) decomposed into total, direct and indirect effects; and C) the proportion of inequalities in COVID-19 deaths mediated by vaccination. Analyses adjusted for individual-level demographics, baseline health, prior infection, and other area-level social determinants of health.

The magnitude of association between income and COVID-19 deaths appeared to be slightly larger for those with than without vaccination (**Appendix Table 4**). However, the difference was not statistically-significant (p-values >0.2). Therefore, in the final mediation analyses model, we did not include interaction between exposure (income) and mediator (vaccination).

In the mediation analyses, lower income was associated with an increased hazard of COVID-19 death during waves four&five both directly and indirectly (via mediation by vaccination) (**Figure 4B**; **Appendix Table 5).** For example, the indirect effect aHR of 1.24 [1.24-1.25] means that for individuals in the lowest-income quintile, their vaccination coverage was associated with 1.24 times hazard of COVID-19 death, compared to if they had the same vaccination coverage as the highest-income quintile. The direct effect aHR of 1.22 [0.88-1.68] means that had vaccination coverage been equal between the highest-income vs. lowest-income quintiles, the lowest-income quintile was associated with 1.22 times hazard of COVID-19 death compared to the highest-income quintile. The proportion mediated by vaccination reflects that 57.9% [21.9%-94.0%] of the total increase in the hazard of COVID-19 death between the lowest-income vs. highest-income quintiles was attributed to differences in vaccination by income (**Figure 4C**; **Appendix Table 5).**

### Sensitivity analyses: unmeasured confounding

We found that a moderate-level of unmeasured confounding (E-value: 1.78-1.80) was required to nullify the indirect effect of income (highest-income vs. lowest-income quintiles) on COVID-19 deaths, and a weak to moderate-level of unmeasured confounding was required to nullify the direct effect (E-value = 1.00-1.74) (**Appendix Table 5**).

### Sensitivity analyses: mediating role of area-level essential work and high-density housing during waves four&five

In two separate mediation analysis models and given observed levels of vaccination, we found that 29.8% [12.3%-7.2%] of the total increase in the hazard of COVID-19 death associated with the lowest-income vs. highest-income quintiles was attributed to patterns of area-level essential workers; while 8.7% [-3.4%-20.8%] was attributed patterns of area-level high-density housing (**Appendix Table 6**).

## Discussion

Across five pandemic waves in Ontario, Canada in 2020-2021, inequalities in COVID-19 deaths by income persisted, even after accounting for individual-level demographics and baseline health (including comorbidities), and other area-level SDOH. By the start of wave four in August 2021, vaccination coverage was lower in lower-income neighbourhoods. Over half (57.9%) of the inequalities in COVID-19 deaths between the lowest and highest income neighbourhoods during the later waves could be attributed to the differential vaccination coverage by income. This means that if vaccine equality had been achieved, inequalities in deaths would persist but be reduced. During the later waves, the magnitude of inequality by income was also larger with Omicron-related deaths than Delta-related deaths. Finally, we found a contrasting pattern in COVID-19 diagnoses and deaths in the context of Omicron infections: lower hazard of diagnosis but higher hazard of death in lower-income neighbourhoods.

The persistent inequalities by income reflect similar patterns in studies from other settings that examined changes over time in COVID-19 deaths by socioeconomic factors(9,10). In the UK, age-standardized inequalities in COVID-19 deaths by area-level socioeconomic factors persisted during 2020-2022, but the analysis was not able to account for confounders such as baseline health(9). In addition, our findings of contrasting patterns in diagnoses and deaths in Omicron infections suggest testing gaps by income during later pandemic waves. Testing gaps by income were previously demonstrated with PCR testing early in the pandemic(4,5,34), with emerging evidence to suggest lower use of SARS-CoV-2 antigen tests associated with lower income during later waves(38). Less access to testing can be a barrier to early and effective isolation support among individuals living in lower-income neighbourhoods, which can lead to onward transmission from undiagnosed infections, and therefore more COVID-19 deaths in lower-income neighbourhoods(39,40).

Our findings on the vaccination gap and the role of vaccination strengthen inference from previous findings, by using population-based study design and individual-level data to mitigate potential biases. The vaccination gap by income confirmed findings from self-reported survey data in Canada (41–43); however, by using population-based vaccination records, our study mitigated the potential for recall and selection biases. Our findings on the mediating role of vaccination in COVID-19 death inequalities are similar to findings from the previous ecological study in the US(10). The US study had found that 37.8% of regional inequalities in COVID-19 case fatality by region-level socioeconomic factors could be attributed to regional inequalities in vaccination coverage (10). By using individual-level vaccination and death data among the total population vs. those diagnosed, our study limited potential ecological fallacy and collider bias (e.g., income and severe COVID-19 outcomes both affect likelihoods of being diagnosed; restricting analyses among samples of diagnosed cases could distort the relationship between income and COVID-19 outcomes)(35). Our findings further support the potential of effect modification by vaccination on social inequalities in COVID-19 deaths, similar to findings from the previous ecological study in the UK(19). Smaller magnitude of inequality in COVID-19 deaths by income among those vaccinated than non-vaccinated may arise if individuals who were vaccinated were also more likely to access other prevention and treatment services than those who were not vaccinated regardless of their area-level income (elaborated in **Appendix Text 3**).

By quantifying the role of vaccination as a mediator of COVID-19 inequalities, our findings indicate that closing the vaccination gap with equal coverage could have reduced inequalities in COVID-19 deaths. However, equal vaccination coverage is not sufficient in addressing inequalities in COVID-19 deaths. Equitable vaccination coverage requires higher levels of coverage among those most at risk at the intersection of SARS-CoV-2 exposures, severity, and onward transmission(44–48). In the context of heterogeneity in onward transmission(39,49), equitable vaccination coverage has been shown to reduce epidemics, curb the spread of new strains(45), and save more lives(45,50). The pathways to equitable coverage includes evidence-informed vaccine prioritization and allocation strategies(39,51), especially in the context of finite and time-constrained supply and delivery, but also implementation. The latter include context-specific and tailored implementation strategies that reduce systemic barriers to vaccine access (18,52,53) and improve vaccine acceptability(54,55). Examples include diversifying vaccination services via mobile, walk-in, drive-through and pop-up clinics, offering more flexible clinic hours, and providing culturally and linguistically appropriate clinics across diversity of communities and lived experience(18,52,53). Our study was not designed to evaluate the impact of specific elements of the vaccination programme in Ontario, which included allocation prioritization to neighbourhoods with the highest per-capita rates of COVID-19 and simultaneous community-tailored implementation strategies(51,56–58). However, our findings signal that additional efforts are needed to close the vaccination gap by income and thereby reduce inequalities in COVID-19 deaths. Future efforts to improve vaccine equity is critical to health equity in COVID-19 deaths and other respiratory viruses where vaccines are effective at preventing infection-attributable deaths.

Even if vaccination coverage had been equal, our findings signal that inequalities in COVID-19 deaths by income would have still persisted. Individuals living in lower-income neighbourhoods are more likely to reside in high density housing with multi-generational households and to work in front-facing essential services; all of which conferred an increased risk of SARS-CoV-2 exposure and transmission(59,60). Indeed, our sensitivity analyses revealed area-level essential work measure could explain 29.8% of inequalities in COVID-19 deaths between the lowest and highest income neighbourhoods. Moreover, individuals in lower-income neighbourhoods were overrepresented in racialized and Indigenous communities where experiences of systemic and institutionalized racism, especially in healthcare, persist as barriers to access to care(18,53,61). Adjustment for area-level SDOH and individual-level demographics and baseline health attenuated but did not nullify the magnitude of inequality(3). Thus, residual heightened risk in COVID-19 deaths by lower-income neighbourhoods might reflect a combination of higher exposure risks associated with individual-level occupation and housing conditions that are not fully captured by area-level measures, and poorer access to and quality of prevention and treatment interventions, including testing(2,62,63) and anti-virals(64). Taken together, our study demonstrates that in addition to vaccination programme, other measures are needed to address exposure and transmission risks (e.g., housing support for multi-generation households; paid sick leave and workplace health and safety measures) and improve access to testing and treatment services.

Limitations include our use of a area-level income measure in the absence of individual-level measures, which might result in an underestimation of social inequalities(65). Individuals who do not have provincial health insurance were not captured and if they were more likely to experience socially disadvantage, we would underestimate inequalities. We did not consider time-dependent vaccination status given the high computational requirement of our large dataset. However, we examined the vaccination gap by income by the start of wave five which was similar to that of wave four (**Appendix Table 7**). We did not consider waning of vaccine protection because we aimed to capture patterns of vaccine access rather than vaccine-induced immunity or protection. If individuals in higher-income neighbourhoods were earlier adopters of vaccines than individuals in lower-income neighbourhoods, waning of vaccine protection would more likely occur in higher-income than lower-income neighbourhoods by the start of wave four (a time when booster dose access was limited). This means our estimates might underestimate the mediating role of vaccination. We lacked data on individual-level SDOH (e.g., occupation, housing condition, and racialized and Indigenous identity), and individuals’ exposures related to contact patterns and physical networks and masking -information that could help further explain the relationship between income and COVID-19 deaths. Our estimates of E-values suggested that unmeasured confounders are not likely to nullify the results of the mediating role of vaccination.

In a high-income country, inequalities by income persisted in COVID-19 deaths due in part to inequalities in vaccination coverage. Additional efforts are needed to address vaccination gaps, but also residual risks of COVID-19 shaped by systemic socioeconomic disadvantage.

## Contribution

LW and SM conceptualized the study with inputs from KB. LW developed the conceptual framework and analysis plan. LW and SS conducted the data cleaning and statistical analyses. AC, MK, and SPH provided critical inputs for data cleaning and statistical analyses. SS, KB, SPH, AS, SDB, PCA, JS, HS, BS, JCK, and SM provided critical input into the results interpretation and preparation of the manuscript. LW wrote the manuscript, with review and edits from all co-authors.

## Conflict of interest

All authors have completed the Unified Competing Interest form (available on request from the corresponding author) and declare: no support from any organisation for the submitted work; no financial relationships with any organisations that might have an interest in the submitted work in the previous three years, no other relationships or activities that could appear to have influenced the submitted work.

SDB participates in Health Canada related programming including vaccination, and COVID-19-related clinical work; and serves on the National Institutes of Health-funded data and safety monitoring boards (unpaid).

## Funding

This work was supported by the Canadian Institutes of Health Research (grant no. VR5-172683; VS1-175536; GA1-177697) and the Ontario Ministry of Health (MOH) funding for COVID-19 and Health Equity Projects. This study was also supported by ICES, which is funded by an annual grant from the Ontario Ministry of Health (MOH) and the Ministry of Long-Term Care (MLTC). This study was also supported by the Ontario Health Data Platform (OHDP), a Province of Ontario initiative to support Ontario’s ongoing response to COVID-19 and its related impacts. The opinions, results and conclusions reported in this paper are those of the authors and are independent from the funding sources. No endorsement by the OHDP, its partners, or the Province of Ontario is intended or should be inferred.

Sharmistha Mishra is supported by a Tier 2 Canada Research Chair in Mathematical Modelling and Program Science (CRC-950-232643). Beate Sander is supported by a Tier 2 Canada Research Chair in Economics of Infectious Diseases held by (CRC-950-232429). Janet Smylie is supported by a Tier 1 Canada Research Chair in Advancing Generative Health Services for Indigenous Populations in Canada. Jeffrey Kwong is supported by a Clinician-Scientist Award from the University of Toronto Department of Family and Community Medicine.

## Acknowledgements and disclaimers

This document also used data adapted from the Statistics Canada Postal Code^OM^ Conversion File, which is based on data licensed from Canada Post Corporation, and/or data adapted from the Ontario Ministry of Health Postal Code Conversion File, which contains data copied under license from ©Canada Post Corporation and Statistics Canada. Adapted from Statistics Canada, Canadian Census 2016. This does not constitute an endorsement by Statistics Canada of this product.

Parts of this material are based on data and/or information compiled and provided by: MOH, Canadian Institute for Health Information (CIHI), Cancer Care Ontario (CCO), Immigration, Refugees and Citizenship Canada (IRCC), Statistics Canada, and IQVIA Solutions Canada Inc. The analyses, conclusions, opinions and statements expressed herein are solely those of the authors and do not reflect those of the funding or data sources; no endorsement is intended or should be inferred.

We thank IQVIA Solutions Canada Inc. for use of their Drug Information File. The authors are grateful to the 14.7 million Ontario residents without whom this research would be impossible.

## Data sharing

The dataset from this study is held securely in coded form at ICES. While legal data sharing agreements between ICES and data providers (e.g., healthcare organizations and government) prohibit ICES from making the dataset publicly available, access may be granted to those who meet pre-specified criteria for confidential access, available at www.ices.on.ca/DAS. The full dataset creation plan and underlying analytic code are available from the authors upon request, understanding that the computer programs may rely upon coding templates or macros that are unique to ICES and are therefore either inaccessible or may require modification.

## Notes

### Author Declarations

Data use is authorized under section 45 of Ontario Personal Health Information Protection Act and does not require review by a Research Ethics Board.

## References

1. WHO Coronavirus (COVID-19) Dashboard [Internet]. [cited 2023 Sep 15]. Available from: https://covid19.who.int

2. Riou J, Panczak R, Althaus CL, Junker C, Perisa D, Schneider K, et al. Socioeconomic position and the COVID-19 care cascade from testing to mortality in Switzerland: a population-based analysis. Lancet Public Health. 2021 Sep;6(9):e683–91.

3. Wang L, Calzavara A, Baral S, Smylie J, Chan AK, Sander B, et al. Differential Patterns by Area-level Social Determinants of Health in Coronavirus Disease 2019 (COVID-19)–related Mortality and Non– COVID-19 Mortality: A Population-based Study of 11.8 Million People in Ontario, Canada. Clin Infect Dis. 2022 Oct 28;ciac850.

4. Williamson EJ, Walker AJ, Bhaskaran K, Bacon S, Bates C, Morton CE, et al. OpenSAFELY: factors associated with COVID-19 death in 17 million patients. Nature. 2020 Aug 1;584(7821):430–6.

5. Karmakar M, Lantz PM, Tipirneni R. Association of Social and Demographic Factors With COVID-19 Incidence and Death Rates in the US. JAMA Netw Open. 2021 Jan 29;4(1):e2036462.

6. Mena GE, Martinez PP, Mahmud AS, Marquet PA, Buckee CO, Santillana M. Socioeconomic status determines COVID-19 incidence and related mortality in Santiago, Chile. Science. 2021 May 28;372(6545):eabg5298.

7. Arceo-Gomez EO, Campos-Vazquez RM, Esquivel G, Alcaraz E, Martinez LA, Lopez NG. The income gradient in COVID-19 mortality and hospitalisation: An observational study with social security administrative records in Mexico. Lancet Reg Health - Am. 2021 Nov 10;6:100115.

8. Hussey H, Zinyakatira N, Morden E, Ismail M, Paleker M, Bam JL, et al. Higher COVID-19 mortality in low-income communities in the City of Cape Town – a descriptive ecological study. Gates Open Res. 2021 Jun 4;5:90.

9. The continuing impact of COVID-19 on health and inequalities - The Health Foundation [Internet]. [cited 2023 Sep 21]. Available from: https://www.health.org.uk/publications/long-reads/the-continuing-impact-of-covid-19-on-health-and-inequalities

10. Chen Y, Zhang L, Li T, Li L. Amplified effect of social vulnerability on health inequality regarding COVID-19 mortality in the USA: the mediating role of vaccination allocation. BMC Public Health. 2022 Nov 19;22(1):2131.

11. Ma H, Chan AK, Baral SD, Fahim C, Straus S, Sander B, et al. Which Curve Are We Flattening? The Disproportionate Impact of COVID-19 Among Economically Marginalized Communities in Ontario, Canada, Was Unchanged From Wild-Type to Omicron. Open Forum Infect Dis. 2023 Jan 1;10(1):ofac690.

12. Watson OJ, Barnsley G, Toor J, Hogan AB, Winskill P, Ghani AC. Global impact of the first year of COVID-19 vaccination: a mathematical modelling study. Lancet Infect Dis. 2022 Sep 1;22(9):1293–302.

13. Bayati M, Noroozi R, Ghanbari-Jahromi M, Jalali FS. Inequality in the distribution of Covid-19 vaccine: a systematic review. Int J Equity Health. 2022 Aug 30;21(1):122.

14. Roghani A. The relationship between macro-socioeconomics determinants and COVID-19 vaccine distribution. AIMS Public Health. 2021;8(4):655–64.

15. Oxfam International [Internet]. 2022 [cited 2023 Sep 17]. Pandemic of greed. Available from: https://www.oxfam.org/en/research/pandemic-greed

16. Brookings [Internet]. [cited 2024 Jan 3]. Why global vaccine equity is the prescription for a full recovery. Available from: https://www.brookings.edu/articles/why-global-vaccine-equity-is-the-prescription-for-a-full-recovery/

17. MacDonald NE, Comeau J, Dubé È, Graham J, Greenwood M, Harmon S, et al. Royal society of Canada COVID-19 report: Enhancing COVID-19 vaccine acceptance in Canada. FACETS. 2021 Jan;6:1184–246.

18. Marfo EA, Aylsworth L, Driedger SM, MacDonald SE, Manca T. The Conversation. 2022 [cited 2023 Nov 21]. Beyond vaccine hesitancy: Understanding systemic barriers to getting vaccinated. Available from: http://theconversation.com/beyond-vaccine-hesitancy-understanding-systemic-barriers-to-getting-vaccinated-193610

19. Sá F. Do vaccinations reduce inequality in Covid-19 mortality? Evidence from England. Soc Sci Med. 2022 Jul;305:115072.

20. Public Health Ontario. COVID-19 in Ontario: Focus on August 28, 2022 to September 3, 2022 (Week 35). 2022 [cited 2023 Nov 21]; Available from: https://www.publichealthontario.ca/-/media/Documents/nCoV/epi/2022/09/weekly-epi-summary-covid-ontario-sept-8.pdf?rev=8b9072357fc1402094f347deffe7a816&la=fr

21. Dictionary, Census of Population, 2016 - Dissemination area (DA) [Internet]. [cited 2023 Sep 18]. Available from: https://www12.statcan.gc.ca/census-recensement/2016/ref/dict/geo021-eng.cfm

22. Wachtler B, Michalski N, Nowossadeck E, Diercke M, Wahrendorf M, Santos-Hövener C, et al. Socioeconomic inequalities and COVID-19 – A review of the current international literature. J Health Monit. 2020 Oct 9;5(Suppl 7):3–17.

23. Chiu M, Lebenbaum M, Lam K, Chong N, Azimaee M, Iron K, et al. Describing the linkages of the immigration, refugees and citizenship Canada permanent resident data and vital statistics death registry to Ontario’s administrative health database. BMC Med Inform Decis Mak. 2016 Oct 21;16(1):135.

24. Permanent Resident Data (Immigration, Refugees and Citizenship Canada) - Ontario Data Catalogue [Internet]. [cited 2024 Jan 11]. Available from: https://data.ontario.ca/dataset/permanent-resident-data-immigration-refugees-and-citizenship-canada

25. studylib.net [Internet]. [cited 2023 Nov 14]. The Johns Hopkins ACG® System, Technical Reference Guide. Available from: https://studylib.net/doc/8365333/the-johns-hopkins-acg®-system--technical-reference-guide

26. ICES [Internet]. [cited 2023 Sep 18]. ICES | Use ICES Data | Working With ICES Data. Available from: https://www.ices.on.ca/use-ices-data/working-with-ices-data/

27. Austin PC, Lee DS, Fine JP. Introduction to the Analysis of Survival Data in the Presence of Competing Risks. Circulation. 2016 Feb 9;133(6):601–9.

28. Lau B, Cole SR, Gange SJ. Competing Risk Regression Models for Epidemiologic Data. Am J Epidemiol. 2009 Jul 15;170(2):244–56.

29. SAS Institute, Inc. SAS/ACCESS® 9.4 Interface to ADABAS. Cary, NC: SAS Institute, Inc, 2013.

30. Lin DY, Wei LJ. The Robust Inference for the Cox Proportional Hazards Model. J Am Stat Assoc. 1989;84(408):1074–8.

31. Harrigan SP. The mediating role of SARS-CoV-2 variants between income and hospitalisation due to COVID-19: A period-based mediation analysis. Under review. AJE-00910-2023.R1.

32. Valeri L, VanderWeele TJ. Mediation analysis allowing for exposure-mediator interactions and causal interpretation: theoretical assumptions and implementation with SAS and SPSS macros. Psychol Methods. 2013 Jun;18(2):137–50.

33. VanderWeele TJ, Robinson WR. On the Causal Interpretation of Race in Regressions Adjusting for Confounding and Mediating Variables: Epidemiology. 2014 Jul;25(4):473–84.

34. R Core Team. R: a language and environment for statistical computing. [Internet]. Vienna, Austria: R Foundation for Statistical Computing; 2022 [cited 2023 Nov 21]. Available from: https://www.r-project.org/

35. Shi B, Choirat C, Coull BA, VanderWeele TJ, Valeri L. CMAverse: A Suite of Functions for Reproducible Causal Mediation Analyses. Epidemiology. 2021 Sep;32(5):e20.

36. VanderWeele TJ, Ding P. Sensitivity Analysis in Observational Research: Introducing the E-Value. Ann Intern Med. 2017 Aug 15;167(4):268.

37. Smith LH, VanderWeele TJ. Mediational E-values: Approximate Sensitivity Analysis for Unmeasured Mediator-Outcome Confounding. Epidemiol Camb Mass. 2019 Nov;30(6):835–7.

38. Rader B, Gertz A, Iuliano AD, Gilmer M, Wronski L, Astley CM, et al. Use of At-Home COVID-19 Tests — United States, August 23, 2021–March 12, 2022. Morb Mortal Wkly Rep. 2022 Apr 1;71(13):489–94.

39. Mishra S, Kwong JC, Chan AK, Baral SD. Understanding heterogeneity to inform the public health response to COVID-19 in Canada. CMAJ. 2020 Jun 22;192(25):E684–5.

40. Kucharski AJ, Klepac P, Conlan AJK, Kissler SM, Tang ML, Fry H, et al. Effectiveness of isolation, testing, contact tracing, and physical distancing on reducing transmission of SARS-CoV-2 in different settings: a mathematical modelling study. Lancet Infect Dis. 2020 Oct;20(10):1151–60.

41. Public Health Agency of Canada. COVID-19 vaccine uptake and intent: Canadian Community Health Survey (CCHS) insight [Internet]. 2021 [cited 2023 Sep 20]. Available from: https://www.canada.ca/en/public-health/services/publications/vaccines-immunization/covid-19-vaccine-uptake-intent-canadian-community-health-survey.html

42. Public Health Agency of Canada. COVID-19 Vaccination Coverage Survey (CVCS): Cycle 2 full report [Internet]. 2021 [cited 2023 Nov 21]. Available from: https://www.canada.ca/en/public-health/services/publications/vaccines-immunization/covid-19-vaccination-coverage-survey/full-report-cycle-2.html

43. Ortiz AM, Nasri B. Socio-demographic determinants of COVID-19 vaccine uptake in Ontario: Exploring differences across the Health Region model [Internet]. medRxiv; 2023 [cited 2023 Nov 21]. p. 2023.08.04.23293662. Available from: https://www.medrxiv.org/content/10.1101/2023.08.04.23293662v1

44. UNDP [Internet]. [cited 2024 Jan 3]. Global dashboard for vaccine equity | Data Futures Platform. Available from: https://data.undp.org/insights/vaccine-equity

45. Ye Y, Zhang Q, Wei X, Cao Z, Yuan HY, Zeng DD. Equitable access to COVID-19 vaccines makes a life-saving difference to all countries. Nat Hum Behav. 2022 Feb;6(2):207–16.

46. Beste LA, Chen A, Geyer J, Wilson M, Schuttner L, Wheat C, et al. Best Practices for an Equitable Covid-19 Vaccination Program. Nejm Catal Innov Care Deliv. 2021 Sep 15;2(10):10.1056/CAT.21.0238.

47. World Health Organization. Accelerating COVID-19 Vaccine Deployment. Removing obstacles to increase coverage levels and protect those at high risk [Internet]. [cited 2024 Jan 3]. Available from: https://www.who.int/publications/m/item/accelerating-covid-19-vaccine-deployment

48. Cevik M, Baral SD. Networks of SARS-CoV-2 transmission. Science. 2021 Jul 9;373(6551):162–3.

49. Wang L, Ma H, Yiu KCY, Calzavara A, Landsman D, Luong L, et al. Heterogeneity in testing, diagnosis and outcome in SARS-CoV-2 infection across outbreak settings in the Greater Toronto Area, Canada: an observational study. Can Med Assoc Open Access J. 2020 Oct 1;8(4):E627–36.

50. Agrawal V, Sood N, Whaley CM. The Impact of the Global COVID-19 Vaccination Campaign on All-Cause Mortality [Internet]. National Bureau of Economic Research; 2023 [cited 2023 Nov 9]. (Working Paper Series). Available from: https://www.nber.org/papers/w31812

51. Ontario COVID-19 Science Advisory Table [Internet]. [cited 2023 Sep 20]. A Vaccination Strategy for Ontario COVID-19 Hotspots and Essential Workers. Available from: https://covid19-sciencetable.ca/sciencebrief/a-vaccination-strategy-for-ontario-covid-19-hotspots-and-essential-workers/

52. Aylsworth L, Manca T, Dubé È, Labbé F, Driedger SM, Benzies K, et al. A qualitative investigation of facilitators and barriers to accessing COVID-19 vaccines among Racialized and Indigenous Peoples in Canada. Hum Vaccines Immunother. 2022 Nov 30;18(6):2129827.

53. Smylie J, McConkey S, Rachlis B, Avery L, Mecredy G, Brar R, et al. Uncovering SARS-COV-2 vaccine uptake and COVID-19 impacts among First Nations, Inuit and Métis Peoples living in Toronto and London, Ontario. CMAJ. 2022 Aug 2;194(29):E1018–26.

54. Cénat JM, Noorishad PG, Bakombo SM, Onesi O, Mesbahi A, Darius WP, et al. A Systematic Review on Vaccine Hesitancy in Black Communities in Canada: Critical Issues and Research Failures. Vaccines. 2022 Nov;10(11):1937.

55. Lavoie K, Gosselin-Boucher V, Stojanovic J, Gupta S, Gagné M, Joyal-Desmarais K, et al. Understanding national trends in COVID-19 vaccine hesitancy in Canada: results from five sequential cross-sectional representative surveys spanning April 2020–March 2021. BMJ Open. 2022 Apr 1;12(4):e059411.

56. Office of the Auditor General of Ontario. Value-for-Money Audit: COVID-19 Vaccination Program [Internet]. 2022 [cited 2023 Nov 21]. Available from: https://www.auditor.on.ca/en/content/annualreports/arreports/en22/AR_COVIDVaccination_en22.pdf

57. Toronto C of. City of Toronto. City of Toronto; 2022 [cited 2024 Jan 3]. COVID-19: Vaccine Engagement Teams Updates. Available from: https://www.toronto.ca/community-people/health-wellness-care/health-programs-advice/respiratory-viruses/covid-19/covid-19-vaccines/covid-19-city-immunization-program/vaccine-engagement-teams-updates/

58. acymbalista. Durham Health Department offering pop-up vaccination clinics in local schools [Internet]. PVNCCDSB. 2021 [cited 2024 Jan 3]. Available from: https://www.pvnccdsb.on.ca/durham-health-department-offering-pop-up-vaccination-clinics-in-local-schools/

59. Ahmad K, Erqou S, Shah N, Nazir U, Morrison AR, Choudhary G, et al. Association of poor housing conditions with COVID-19 incidence and mortality across US counties. PLoS ONE [Internet]. 2020 [cited 2023 Nov 21];15(11). Available from: https://www.ncbi.nlm.nih.gov/pmc/articles/PMC7605696/

60. Rao A, Ma H, Moloney G, Kwong JC, Jüni P, Sander B, et al. A disproportionate epidemic: COVID-19 cases and deaths among essential workers in Toronto, Canada. Ann Epidemiol. 2021 Nov;63:63–7.

61. Government of Canada SC. Income statistics by Indigenous identity and residence by Indigenous geography: Canada, provinces and territories [Internet]. 2022 [cited 2024 Jan 11]. Available from: https://www150.statcan.gc.ca/t1/tbl1/en/tv.action?pid=9810028101

62. Vandentorren S, Smaïli S, Chatignoux E, Maurel M, Alleaume C, Neufcourt L, et al. The effect of social deprivation on the dynamic of SARS-CoV-2 infection in France: a population-based analysis. Lancet Public Health. 2022 Mar 1;7(3):e240–9.

63. O’Neill B, Kalia S, Hum S, Gill P, Greiver M, Kirubarajan A, et al. Socioeconomic and immigration status and COVID-19 testing in Toronto, Ontario: retrospective cross-sectional study. BMC Public Health. 2022 May 29;22(1):1067.

64. Shaikh TG, Waseem S, Ahmed SH, Asghar MS, Tahir MJ. Disproportionate distribution of coronavirus disease 2019 (COVID-19) antiviral pills: Vaccine inequity replay? Infect Control Hosp Epidemiol.:1–2.

65. Moss JL, Johnson NJ, Yu M, Altekruse SF, Cronin KA. Comparisons of individual- and area-level socioeconomic status as proxies for individual-level measures: evidence from the Mortality Disparities in American Communities study. Popul Health Metr. 2021 Jan 7;19(1):1.

