## Appendix for "Social inequalities in COVID-19 deaths by area-level income: patterns over time and the mediating role of vaccination in a population of 11.2 million people in Ontario, Canada"

**Appendix Table 1.** Patterns in COVID-19 deaths and diagnoses by area-level income across pandemic waves.

|  | COVID-19 death^a^ | | COVID-19 diagnosis | |
| --- | --- | --- | --- | --- |
| Area-level income stratified by wave^b^ | Hazard ratio^c^ (95% CI) | | Hazard ratio^c^(95% CI) | |
|  | Unadjusted | Adjusted^d^ | Unadjusted | Adjusted^d^ |
| Wave 1: Income (5= Highest)^e^ |  |  |  |  |
| 4 vs 5 | 1.04 (0.82, 1.32) | 1.14 (0.89, 1.46) | 1.31 (1.24, 1.38) | 0.94 (0.90, 0.98) |
| 3 vs 5 | 1.43 (1.14, 1.78) | 1.34 (1.04, 1.72) | 1.58 (1.50, 1.66) | 0.97 (0.93, 1.02) |
| 2 vs 5 | 1.65 (1.33, 2.04) | 1.26 (0.95, 1.67) | 2.22 (2.11, 2.33) | 0.86 (0.81, 0.90) |
| 1 vs 5 | 2.07 (1.68, 2.54) | 1.37 (0.98, 1.92) | 3.73 (3.57, 3.91) | 0.87 (0.81, 0.92) |
| Wave 2:Income (5= Highest)^e^ |  |  |  |  |
| 4 vs 5 | 1.07 (0.93, 1.23) | 1.05 (0.91, 1.22) | 1.27 (1.25, 1.30) | 1.02 (1.00, 1.04) |
| 3 vs 5 | 1.45 (1.27, 1.65) | 1.17 (1.00, 1.36) | 1.79 (1.75, 1.83) | 1.06 (1.04, 1.08) |
| 2 vs 5 | 1.93 (1.70, 2.18) | 1.22 (1.03, 1.45) | 2.54 (2.49, 2.60) | 1.04 (1.01, 1.06) |
| 1 vs 5 | 2.29 (2.02, 2.58) | 1.21 (0.99, 1.48) | 4.53 (4.44, 4.61) | 1.03 (1.00, 1.05) |
| Wave 3:Income (5= Highest)^e^ |  |  |  |  |
| 4 vs 5 | 1.31 (1.10, 1.56) | 1.11 (0.93, 1.33) | 1.26 (1.23, 1.29) | 1.07 (1.05, 1.09) |
| 3 vs 5 | 1.88 (1.60, 2.21) | 1.26 (1.04, 1.51) | 1.74 (1.70, 1.78) | 1.11 (1.08, 1.13) |
| 2 vs 5 | 2.24 (1.91, 2.62) | 1.21 (0.98, 1.48) | 2.48 (2.43, 2.53) | 1.09 (1.07, 1.12) |
| 1 vs 5 | 3.42 (2.95, 3.97) | 1.55 (1.22, 1.96) | 3.94 (3.86, 4.01) | 1.14 (1.11, 1.17) |
| Wave 4&5:Income (5= Highest)^e^ |  |  |  |  |
| 4 vs 5 | 1.24 (0.99, 1.56) | 1.17 (0.92, 1.48) | 1.20 (1.18, 1.22) | 0.98 (0.97, 1.00) |
| 3 vs 5 | 1.49 (1.20, 1.85) | 1.31 (1.02, 1.68) | 1.41 (1.38, 1.43) | 0.94 (0.93, 0.96) |
| 2 vs 5 | 1.99 (1.62, 2.45) | 1.53 (1.17, 2.01) | 1.58 (1.56, 1.60) | 0.89 (0.87, 0.91) |
| 1 vs 5 | 2.20 (1.79, 2.70) | 1.57 (1.15, 2.15) | 1.47 (1.45, 1.49) | 0.86 (0.84, 0.88) |
| Trend test^f^(p-values) |  |  |  |  |
| 4 vs 5 | NA | 0.089 | NA | NA |
| 3 vs 5 | NA | 0.17 | NA | NA |
| 2 vs 5 | NA | 0.081 | NA | NA |
| 1 vs 5 | NA | 0.02 | NA | NA |

^a^Death within 30 days following or 7 days prior to a lab-confirmed positive SARS-CoV-2 test was considered a COVID-19 death.

^b^Waves were defined based on the date of diagnosis and classified as follows: wave 1: Mar 1-Aug 31, 2020; wave 2: Sep 1, 2020–Feb 28, 2021; wave 3: Mar 1–Jul 31, 2021; waves 4&5: Aug 1-Dec 31, 2021.

^c^Cause-specific hazard models were used to examine COVID-19 deaths (treating deaths not related to COVID-19 as competing risk events) and to examine COVID-19 diagnosis (treating all deaths as competing risk events).

^d^Adjusted for individual-level demographics (age, sex, immigration status, residing in rural vs. urban, public health unit of residence), baseline health (comorbidity index measured by the Aggregated Diagnosis Groups, past hospitalization and outpatient visits), and other area-level social determinants of health (educational attainment, essential workers, ethnic diversity and housing condition).

^e^Dissemination areas were ranked within each city by their median household income to create income quintiles; we ranked within city instead of within the entire province to take the cost of living into account; a disseminaton area being in quintile 1 means it is among the lowest 20% of dissenmination areas in its city by median household income.

^f^Trend tests were performed by fitting an additonal model using the full sample across all pandemic waves, adding an interaction term between area-level income (treated as a categegorical variable) and pandemic waves (treated as a continuous variable), to examine the trend over time in the magnitude of association between income and COVID-19 deaths.

Abbreviations: CI, confidence interval; NA: not applicable because analyses were not performed.

**Appendix Table 2.** Patterns in COVID-19 variant-specific deaths and diagnoses by area-level income during waves four and five (diagnosis date between Aug 1-Dec 31, 2021).

|  | Delta-related^b^ outcome | | Omicron-related^b^ outcome | |
| --- | --- | --- | --- | --- |
| Area-level income^a^ | Hazard ratio^c^ (95% CI) | | Hazard ratio^c^ (95% CI) | |
|  | Unadjusted | Adjusted^d^ | Unadjusted | Adjusted^d^ |
| Outcome: variant-specific death^e^ |  |  |  |  |
| Income (5= Highest) |  |  |  |  |
| 4 vs 5 | 1.21 (0.91, 1.61) | 1.02 (0.76, 1.37) | 1.30 (0.86, 1.97) | 1.53 (1.02, 2.30) |
| 3 vs 5 | 1.42 (1.08, 1.87) | 1.08 (0.79, 1.48) | 1.67 (1.13, 2.48) | 1.95 (1.26, 3.01) |
| 2 vs 5 | 1.86 (1.43, 2.41) | 1.20 (0.85, 1.69) | 2.15 (1.48, 3.13) | 2.33 (1.46, 3.73) |
| 1 vs 5 | 2.22 (1.72, 2.87) | 1.29 (0.87, 1.92) | 2.04 (1.39, 2.98) | 2.24 (1.28, 3.93) |
| Outcome: variant-specific diagnosis |  |  |  |  |
| Income (5= Highest) |  |  |  |  |
| 4 vs 5 | 1.08 (1.05, 1.11) | 1.03 (1.00, 1.06) | 0.96 (0.95, 0.98) | 0.97 (0.95, 0.98) |
| 3 vs 5 | 1.10 (1.07, 1.13) | 1.03 (1.00, 1.07) | 0.90 (0.88, 0.91) | 0.92 (0.91, 0.94) |
| 2 vs 5 | 1.09 (1.06, 1.12) | 1.01 (0.98, 1.05) | 0.81 (0.80, 0.83) | 0.85 (0.83, 0.87) |
| 1 vs 5 | 1.19 (1.16, 1.23) | 1.08 (1.03, 1.13) | 0.75 (0.74, 0.77) | 0.79 (0.77, 0.81) |

^a^Dissemination areas were ranked within each city by their median household income to create income quintiles; we ranked within city instead of within the entire provinceto to take cost of living into account; a disseminaton area being in quintile 1 means it is among the lowest 20% of dissenmination areas in its city by median household income.

^b^Classification of COVID-19 variants are detailed in **Appendix text 2**.

^c^Cause-specific hazard models were used to examine COVID-19 variant-specific death (treating deaths not related to COVID-19, and deaths related to other variants as competing risk events) and to examine COVID-19 variant-specific diagnosis (treating all deaths and diagnosis of other variants as competing risk events).

^d^Adjusted for individual-level demographics (age, sex, immigration status, residing in rural vs. urban, public health unit of residence), baseline health (comorbidity index measured by the Aggregated Diagnosis Groups, past hospitalization and outpatient visits), and other area-level social determinants of health (educational attainment, essential workers, ethnic diversity and housing condition).

^e^Death within 30 days following or 7 days prior to a lab-confirmed positive SARS-CoV-2 test was considered a COVID-19 death; COVID-19 death with a diagnosis with the Delta variant is classified as a Delta-related death; COVID-19 death with a diagnosis with the Omicron variant is classified as an Omicron-related death.

Abbreviations: CI, confidence interval.

**Appendix Table 3.** Patterns in vaccination by the start of wave four and in COVID-19 deaths during waves four and five (diagnosis date between Aug 1-Dec 31, 2021).

|  | Vaccination status^b^ | | COVID-19 death^c^ | |
| --- | --- | --- | --- | --- |
| Area-level Income^a^ | Odds ratio^d^ (95% CI) | | Hazard ratio^e^(95% CI) | |
|  | Unadjusted | Adjusted^f^ | Unadjusted | Adjusted^g^ |
| Income (5= Highest) |  |  |  |  |
| 4 vs 5 | 0.80 (0.80, 0.81) | 0.90 (0.90, 0.91) | 1.09 (0.87, 1.37) | 1.07 (0.84, 1.36) |
| 3 vs 5 | 0.71 (0.71, 0.72) | 0.86 (0.86, 0.87) | 1.24 (0.99, 1.54) | 1.18 (0.92, 1.51) |
| 2 vs 5 | 0.64 (0.64, 0.64) | 0.82 (0.81, 0.82) | 1.56 (1.27, 1.92) | 1.31 (1.00, 1.72) |
| 1 vs 5 | 0.50 (0.50, 0.51) | 0.71 (0.70, 0.71) | 1.52 (1.24, 1.87) | 1.22 (0.88, 1.68) |

^a^Dissemination areas were ranked within each city by their median household income to create income quintiles; we ranked within city instead of within the entire province to take the cost of living into account; a disseminaton area being in quintile 1 means it is among the lowest 20% of dissenmination areas in its city by median household income.

^b^Vaccination status defined as receiving ≥ 1 dose of Johnson-Johnson vaccine or ≥ 2 doses of other vaccines. Details in Appendix text 1.

^c^Death within 30 days following or 7 days prior to a lab-confirmed positive SARS-CoV-2 test was considered a COVID-19 death.

^d^Logistic regression models were used to examine the odds of vaccination.

^e^Cause-specific hazard models were used to examine COVID-19 death (treating deaths not related to COVID-19 as competing risk events).

^f^Adjusted for individual-level demographics (age, sex, immigration status, residing in rural vs. urban, public health unit of residence), baseline health (comorbidity index measured by the Aggregated Diagnosis Groups, past hospitalization and outpatient visits), prior infection, and other area-level social determinants of health (educational attainment, essential workers, ethnic diversity and housing condition).

^g^Adjusted for the list of covariates in footnote f, and vaccination status (yes/no) by start of wave 4.

Abbreviations: CI, confidence interval.

**Appendix Table 4.** Patterns in COVID-19 deaths^a^ during waves four and five by area-level income^b^ stratified by vaccination status^c^.

|  | Fully vacciated individuals | | Not fully vaccinated individuals | | P-values (for the interaction term between income and vaccnation) | |
| --- | --- | --- | --- | --- | --- | --- |
| Area-level Income | Hazard ratio^d^(95% CI) | | Hazard ratio^d^(95% CI) | |  |  |
|  | Unadjusted | Adjusted^e^ | Unadjusted | Adjusted^e^ | Unadjusted | Adjusted^e^ |
| Income (5= Highest) |  |  |  |  |  |  |
| 4 vs 5 | 1.08 (0.78, 1.49) | 1.08 (0.78, 1.51) | 1.30 (0.94, 1.80) | 1.16 (0.83, 1.62) | 0.46 | 0.43 |
| 3 vs 5 | 1.28 (0.94, 1.75) | 1.17 (0.81, 1.68) | 1.49 (1.09, 2.04) | 1.30 (0.92, 1.83) | 0.58 | 0.70 |
| 2 vs 5 | 1.55 (1.14, 2.09) | 1.21 (0.81, 1.79) | 2.03 (1.51, 2.73) | 1.59 (1.09, 2.30) | 0.36 | 0.50 |
| 1 vs 5 | 1.83 (1.36, 2.47) | 1.29 (0.81, 2.06) | 1.96 (1.46, 2.62) | 1.45 (0.95, 2.22) | 0.40 | 0.29 |

^a^Death within 30 days following or 7 days prior to a lab-confirmed positive SARS-CoV-2 test was considered a COVID-19 death.

^b^Dissemination areas were ranked within each city by their median household income to create income quintiles; we ranked within city instead of within the entire province to take the cost of living into account; a disseminaton area being in quintile 1 means it is among the lowest 20% of dissenmination areas in its city by median household income.

^c^Vaccination status defined as receiving ≥ 1 dose of Johnson-Johnson vaccine or ≥ 2 doses of other vaccines. Details in Appendix text 1.

^d^ Estimated using cause-specific hazard models, treating deaths not related to COVID-19 as competing risk events.

^e^Adjusted for individual-level demographics (age, sex, immigration status, residing in rural vs. urban, public health unit of residence), baseline health (comorbidity index measured by the Aggregated Diagnosis Groups, past hospitalization and outpatient visits), prior infection, and other area-level social determinants of health (educational attainment, essential workers, ethnic diversity and housing condition).Abbreviations: CI, confidence interval.

**Appendix Table 5.** Results of causal mediation analyses and sensitivity analyses for unmeasured confounding for the relationship between area-level income^a^ (exposure) and COVID-19 deaths^b^ (outcome) during waves four and five (diagnosis date between Aug 1-Dec 31, 2021), with vaccination status^c^ as the mediator.

|  | **Estimate** | |
| --- | --- | --- |
| **Effect decomposition**^d^ | Adjusted^e^ hazard ratios or proportion mediated % (95% CI) | **E-value (95% LCI)**^f^ |
| Income quintile 4 vs. 5 (highest) |  |  |
| Total effect | 1.15 (0.90, 1.45) | 1.56 (1.00) |
| Direct effect | 1.07 (0.84, 1.36) | 1.36 (1.00) |
| Indirect effect | 1.07 (1.07, 1.07) | 1.34 (1.33) |
| Proportion mediated | 50.9% (-32.8%, 134.6%) | NA |
| Income quintile 3 vs. 5 (highest) |  |  |
| Total effect | 1.30 (1.01, 1.66) | 1.93 (1.13) |
| Direct effect | 1.18 (0.92, 1.51) | 1.65 (1.00) |
| Indirect effect | 1.10 (1.10, 1.11) | 1.43 (1.42) |
| Proportion mediated | 39.8% (6.5%, 73.1%) | NA |
| Income quintile 2 vs. 5 (highest) |  |  |
| Total effect | 1.50 (1.15, 1.97) | 2.38 (1.56) |
| Direct effect | 1.31 (1.00, 1.72) | 1.97 (1.08) |
| Indirect effect | 1.14 (1.14, 1.15) | 1.55 (1.53) |
| Proportion mediated | 37.4% (17.1%, 57.6%) | NA |
| Income quintile 1 vs. 5 (highest) |  |  |
| Total effect | 1.52 (1.10, 2.10) | 2.41 (1.44) |
| Direct effect | 1.22 (0.88, 1.68) | 1.74 (1.00) |
| Indirect effect | 1.25 (1.24, 1.25) | 1.80 (1.78) |
| Proportion mediated | 57.9% (21.9%, 94.0%) | NA |

^a^Dissemination areas were ranked within each city by their median household income to create income quintiles; we ranked within city instead of within the entire province to take the cost of living into account; a disseminaton area being in quintile 1 means it is among the lowest 20% of dissenmination areas in its city by median household income.

^b^Death within 30 days following or 7 days prior to a lab-confirmed positive SARS-CoV-2 test was considered COVID-19 death.

^c^Vaccination status defined as receiving ≥ 1 dose of Johnson-Johnson vaccine or ≥ 2 doses of other vaccines. Details in Appendix text 1.

^d^Regression-based causal mediation analyses were employed to estimate effect decomposition, where the total effect reflects the association between area-level income and COVID-19 death adjusting for confounders; the direct effect reflects the association between area-level income and COVID-19 death holding the vaccination level the same across income; the indirect effect reflects the association between area-level income and COVID-19 death mediated through vaccination status; and the proportion mediated reflects the proportion of the total effect that was mediated through vaccination status.

^e^Adjusted for confounders including individual-level demographics (age, sex, immigration status, residing in rural vs. urban, public health unit of residence), baseline health (comorbidity index measured by the Aggregated Diagnosis Groups, past hospitalization and outpatient visits), prior infection, and other area-level social determinants of health (educational attainment, essential workers, ethnic diversity and housing condition).

^f^E-value reflects the minimum strength of association (measured as an odds ratio) required between an unmeasured confounder with both an exposure and an outcome to fully explain an observed association. E-values from 1.0-1.5 suggest weak unmeasured confounding is required to nullify the observed effect, while ranges from 1.5-3.0 and >3.0 suggest moderate and strong unmeasured confounding are required, respectively.

Abbreviations: CI, confidence interval; LCI, lower confidence interval; NA, not applicable.

**Appendix Table 6.** Results of two separate causal mediation analyses for the relationship between area-level income^a^ (exposure) and COVID-19 deaths^b^ (outcome) during waves four and five (diagnosis date between Aug 1-Dec 31, 2021), with area-level essential worker and housing density measurements each acting as a mediator in each model.

| **Effect decomposition**^c^ | Adjusted^d^ Hazard ratios or proportion mediated (95% CI) | |
| --- | --- | --- |
| Income quintile 1 vs. 5 (highest) | **Area-level essential worker** (quintile 5 vs. 1 (lowest))^e^ | **Area-level housing density**  (quintile 5 vs. 1 (lowest))^e^ **^f^** |
| Total effect |  |  |
| Direct effect | 1.83 (1.34, 2.48) | 1.65 (1.2, 2.27) |
| Indirect effect | 1.58 (1.15, 2.17) | 1.59 (1.16, 2.2) |
| Proportion mediated | 1.16 (1.08, 1.24) | 1.04 (0.99, 1.08) |
|  | 29.8% (12.3%, 47.2%) | 8.7% (-3.4%, 20.8%) |

^a^Dissemination areas were ranked within each city by their median household income to create income quintiles; we ranked within city instead of within the entire province to take the cost of living into account; a disseminaton area being in quintile 1 means it is among the lowest 20% of dissenmination areas in its city by median household income.

^b^Death within 30 days following or 7 days prior to a lab-confirmed positive SARS-CoV-2 test was considered a COVID-19 death.

^c^Regression-based causal mediation analyses were employed to estimate the effect decomposition, where the total effect reflects the association between area-level income and COVID-19 death adjusting for confounders; the direct effect reflects the association between area-level income and COVID-19 death holding the area-level mediator the same across income; the indirect effect reflects the association between area-level income and COVID-19 death mediated through the corresponding area-level mediator; and the proportion mediated reflects the proportion of the total effect that was mediated through the corresponding area-level mediator.

^d^Adjusted for confounders including individual-level demographics (age, sex, immigration status, residing in rural vs. urban, public health unit of residence), baseline health (comorbidity index measured by the Aggregated Diagnosis Groups, past hospitalization and outpatient visits), prior infection, and other area-level social determinants of health (educational attainment, essential workers (only included for the model with housing density as mediator), ethnic diversity and housing condition); we did not adjust for vaccination in the models as vaccination is in the causal pathway between exposure and outcome, as well as in the causal pathway between the mediator of interest and outcome.

^e^Measured at the dissemination area-level; 1st quintile represents 0%–32.5% of working people in the area who self-identified as working in an essential job, including sales, trades, manufacturing, and agriculture; 2^nd^ quintile, 32.5%–42.3% of people; 3^rd^ quintile, 42.3%–49.8% of people; 4^th^ quintile, 50.0%–57.5% of people; and 5^th^ quintile, 57.5%–114.3% of people.

^f^ Measured at the dissemination area-level; 1^st^ category represents 0–2.6% of households are considered high-density housing; 2^nd^ category, 2.7-5.2%; 3^rd^ category, 5.3-8.7%; 4^th^ category, >8.7%; the high frequency of zeros permitted the creation of only 4 categories (the lower 2 quintiles combined); ‘housing density’/‘housing suitability' refers to whether a private household is living in suitable accommodations according to the National Occupancy Standard; that is, whether the dwelling has enough bedrooms for the size and composition of the household. A household is deemed to be living in suitable accommodations (non-high-density housing) if its dwelling has enough bedrooms, as calculated using the National Occupancy Standard.

Abbreviations: CI, confidence interval.

**Appendix Table 7.** Adjusted patterns in vaccination by the start of wave five (December 15, 2021).

|  | Vaccination status^b^ |
| --- | --- |
| Area-level Income^a^ |  |
|  | Adjusted odds ratio^c,d^ (95% CI) |
| Income (5= Highest) |  |
| 4 vs 5 | 0.94 (0.93, 0.94) |
| 3 vs 5 | 0.90 (0.89, 0.90) |
| 2 vs 5 | 0.85 (0.85, 0.86) |
| 1 vs 5 | 0.74 (0.74, 0.75) |

^a^Dissemination areas were ranked within each city by their median household income to create income quintiles; we ranked within city instead of within the entire province to take the cost of living into account; a disseminaton area being in quintile 1 means it is among the lowest 20% of dissenmination areas in its city by median household income.

^b^Vaccination status defined as receiving ≥ 1 dose of Johnson-Johnson vaccine or ≥ 2 doses of other vaccines. Details in Appendix text 1.

^c^Logistic regression models were used to examine the odds of being fully vaccinated.

^d^Adjusted for individual-level demographics (age, sex, immigration status, residing in rural vs. urban, public health unit of residence), baseline health (comorbidity index measured by the Aggregated Diagnosis Groups, past hospitalization and outpatient visits), prior infection, and other area-level social determinants of health (educational attainment, essential workers, ethnic diversity and housing condition).

Abbreviations: CI, confidence interval.

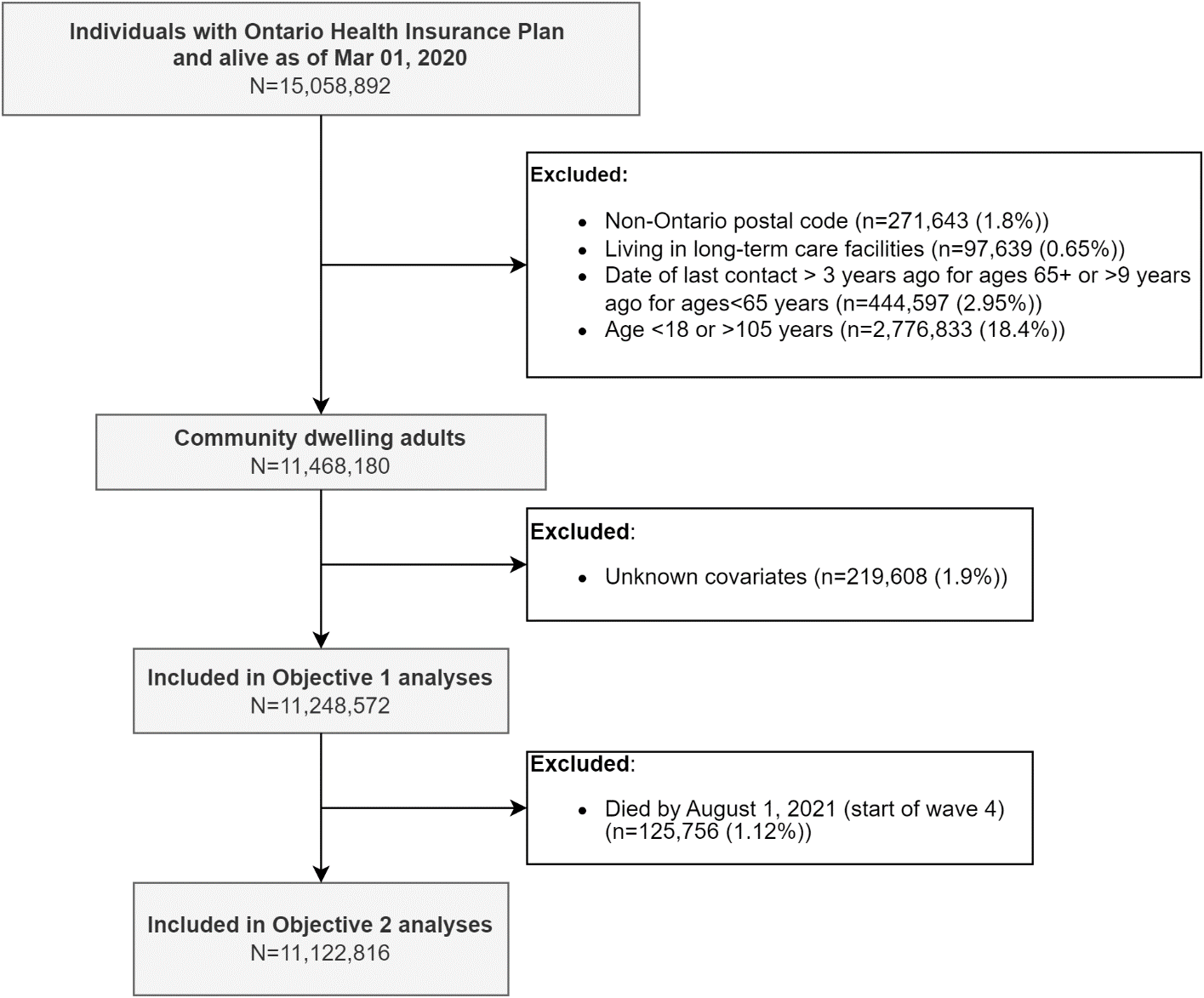

**Appendix Figure 1.** Flow diagram showing inclusion and exclusion criteria and resulting analytic data sets.

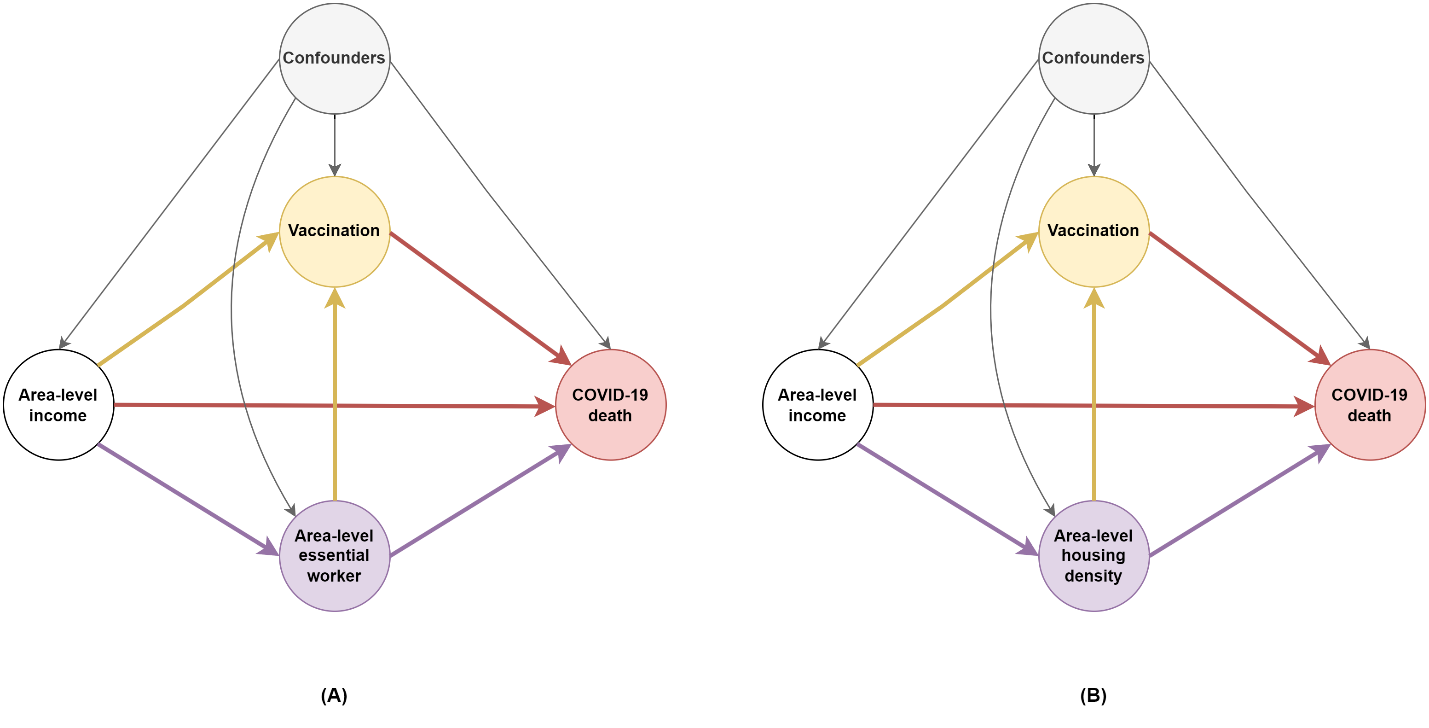

**Appendix Figure 2.** Directed acyclic graphs depicting the relationship between area-level income and COVID-19 deaths as mediated by A) the area-level essential worker measure, and B) the area-level housing density measure. We adjusted for confounders including individual-level demographics, baseline health, prior infection, and other area-level social determinants of health. We did not adjust for vaccination in the models as vaccination is in the causal pathway between exposure and outcome, as well as in the causal pathway between the mediators-of-interest and the outcome.

**Appendix Text 1.** Details on vaccination status classification.

We determined individual’s vaccination status based on Ontario’s COVID-19 vaccination registry. We classified vaccination status into four categories: no vaccination, partially vaccinated, fully vaccinated without booster, and fully vaccinated with booster, based on the date of each vaccine dose in relation to the index date, and the vaccine product type. For the first dose of vaccine, we assume a 14-day lag between the date of vaccine dose administration and the date when vaccine is considered protective; for the second dose of vaccine and onward doses, we assume a 7-day lag between the date of vaccine dose administration and the date when vaccine is considered protective.

Partially vaccinated is defined as i) having one dose of any vaccine other than Janssen Jcovden (Johnson & Johnson) vaccine; or ii) having only two doses of a non-Health Canada-approved vaccine.

Fully vaccinated without booster is defined as i) having two doses of a Health Canada-approved vaccine (Pfizer, Moderna, or Covishield/AstraZeneca); or ii) having one dose of a Janssen Jcovden (Johnson & Johnson) vaccine (1-dose product); or iii) having one dose of a non-Health Canada-approved vaccine and one dose of a Health Canada-approved vaccine (Pfizer, Moderna, Covishield/AstraZeneca).

Fully vaccinated with booster is defined as i) having three doses of any vaccines; or ii) having one dose of Janssen vaccine (1-dose product) plus a booster dose of any other vaccines.

For our regression analyses, we further grouped vaccination status into a binary measure to increase analytic power: fully vaccinated yes vs. no, where fully vaccinated includes fully vaccinated with and without booster.

**Appendix Text 2.** Details on COVID-19 variants’ classification for diagnoses during waves 4&5 (between Aug 1, 2021 and Dec 31, 2021).

In Ontario, the variant testing algorithm has changed over time(1,2). In the period between Aug 1, 2021 and Dec 31, 2021, the following screening criteria were in place: **Aug 1 to Nov 11, 2021:** all polymerase chain reaction (PCR) positive SARS-CoV-2 specimens with cycle threshold (Ct) values ≤ 35 were tested for N501Y mutation and E484K mutation; but positive cases with a N501Y mutation or a E484K mutation detected were **not** reflexed for whole-genome sequencing (WGS); in addition, a proportion of eligible (with Ct≤ 30* from the SARS-CoV-2 PCR diagnostic test and sufficient volume remaining) SARS-CoV-2 positive specimens were selected for WGS; **Nov 12 to Dec 5, 2021:** PCR mutation screening was discontinued. All eligible SARS-CoV-2 positive specimens were sent for WGS; **Dec 6 to Dec 29, 2021:** PCR testing for spike gene target failure (SGTF) was implemented to screen for Omicron for all eligible SARS-CoV-2 positive specimens; the SGTF testing laboratories then submitted a proportion of eligible samples (SGTF PCR Ct≤30 and sufficient volume remaining) for WGS; **Dec 30, 2021:** SGTF testing of all eligible samples was discontinued in Ontario. A proportion of eligible samples were sent for WGS; December 31, 2021: diagnostic PCR testing was restricted to high-risk populations.

We classified COVID-19 variants based on a combination of the following and in the order listed: WGS data when available and conclusive; mutation screening results when available and conclusive; mutation screening results combined with date of diagnosis; and imputation using date of diagnosis only (details in the table below).

| Variants | WGS or mutation screening results | Date range |
| --- | --- | --- |
| Delta | LINEAGE B.1.617 | Not applicable (NA) |
| Delta | LINEAGE B.1.617.2 | NA |
| Delta | LINEAGE AY.3 | NA |
| Delta | MUTATION L452R+ | NA |
| Delta | NO S-GENE TARGET FAILURE | NA |
| Delta | MUTATION N501Y- AND E484K- | All cases with N501Y- AND E484K- were considered as Delta in the period of waves 4&5 (Aug 1-Dec 31, 2021). |
| Delta (imputed) | Missing or inconclusive | For cases tested positive between ’2021-08-15’ and ’2021-11-27’ because among cases with variants known, 98% were Delta during this period in our dataset. |
| Omicron | LINEAGE B.1.1.529 | NA |
| Omicron | S-GENE TARGET FAILURE | For positive tests after Nov 22^nd^ 2021 (first reported Omicron case in Ontario) |
| Omicron | MUTATION N501Y+ AND E484K- | For positive tests after Nov 22^nd^ 2021 (first reported Omicron case in Ontario) |
| Omicron | MUTATION N501Y+ | For positive tests after Nov 22^nd^ 2021 (first reported Omicron case in Ontario) |
| Omicron (imputed) | Missing or inconclusive | For cases tested positive between ’2021-12-21 and ’2021-12-31’ because among cases with variants known, 93.7% were Omicron during this period in our dataset. |

**Appendix text 3.** Hypothesis and interpretations regarding effect modification by vaccination on the relationship between area-level income and COVID-19 deaths.

Hypothesis: the magnitude of inequality in COVID-19 deaths may differ between subgroups with different vaccination status; such that the magnitude of inequality may be smaller in subgroups with vaccination than without vaccination.

Interpretation: In the context of imperfect vaccine protection and potential ‘degree-type’ vaccine protection (vaccine-induced protection reduces infection rates by a fraction on a per-exposure basis), effect modification by vaccination could occur if any of the following (A-C) is true. However, B) is probably the most possible explanation.

- A) Biologically, if vaccination’s protection against severity is dominant and overrides inequality in access and quality of care (e.g., as long as one is vaccinated, infection will not lead to death regardless of access and quality of care);
- B) If those who are vaccinated, are also more likely to access care (including testing and treatment), regardless of their area-level income;
- C) If those who are vaccinated had higher baseline exposure risk, regardless of their area-level income.

Additional notes on hypothesis: in the context of vaccination’s network-level impact (interference) in addition to individual-level impact, and in the context of contact-pattern correlation by area-level income, it maybe counterintuitive to evaluate the social inequality patterns among subgroups with and without vaccination separately as patterns in one group likely impact patterns in the other. In contrast, the role of vaccination as a mediator subsumes unobserved effects of vaccination on individuals who were not vaccinated to fully capture the population-level impact of the vaccine gap on inequalities in COVID-19 deaths.
